# A prescriptive optimization approach to identify minimal barriers to discharge for surgical patients

**DOI:** 10.1101/2023.03.24.23287694

**Authors:** Taghi Khaniyev, Martin S. Copenhaver, Ana Cecilia Zenteno Langle, Kyan C. Safavi, Keren S. Starobinski, Bethany J. Daily, Peter F. Dunn, Retsef Levi

## Abstract

Ensuring timely patient discharges is crucial to managing a hospital’s patient flow; however, discharges are dependent on the coordination of multiple care teams and thus are highly decentralized in nature. Central capacity teams often lack transparency into how to prioritize scarce resources for patients who need them to leave the hospital (e.g., imaging or post-acute facility placement). Our goal is to identify a small subset of so-called barriers to discharge for hospitalized surgical patients by balancing two aims: a high likelihood that the patient will be discharged from the hospital in the next 24 hours if these barriers are resolved; and a high likelihood that these barriers will indeed be resolved. To do so, we combine discharge and barrier-resolution predictive models with a mixed-integer prescriptive optimization model to identify each patient’s minimal barriers. We empirically demonstrate the efficacy of the proposed formulation and solution methodology using data from a large academic medical center. Furthermore, we study the influence of uncertainty in discharge prediction estimates on the prescribed barriers and introduce a robust optimization variant that is capable of overcoming the shortcomings of the nominal approach. Our approach can significantly enhance the ability of capacity management teams to identify which barriers are most important to address to expedite a patient’s discharge while taking into account their inherent heterogeneity.

## 1 Introduction

Large hospitals often operate at or near maximum capacity which causes significant congestion and delays in patient care. Ensuring smooth patient flow in such settings requires central capacity management teams to maintain a delicate balance between hospital admissions (inflow) and discharges (outflow). The inflow of new patients is a mostly transparent process in that the queue of admissions can be centrally tracked within the electronic health record (EHR). Planned admissions (e.g., elective surgeries or chemotherapy administration) are put in the system in advance, and unscheduled admissions through the emergency department (ED) or hospital-to-hospital transfers require a submission of a bed request to a central admitting department. In contrast, the process of discharging patients from the hospital requires coordination of resources across numerous role groups. As a result, capacity planners do not know which beds are likely to become available to match with their admissions queue until they are freed up thereby slowing the intake of incoming patients. Indeed, frequent delays in the discharge process lead to congestion in critical areas like the ED, intensive care units, and the operating rooms (Copenhaver et al., 2019).

Ensuring patients’ timely discharge is a multifactorial challenge. First, there is a high degree of heterogeneity between patients due to their clinical characteristics as well as nonclinical factors such as the availability of social support. Second, discharge management is often spread across multiple groups of care team members (e.g., physicians, nurses, social workers), who in turn may have different perspectives on discharge readiness. Moreover, as hospitals cope with increasing congestion, capacity managers are asked to help prioritize scarce resources (such as imaging or lab tests) in order to expedite patient flow. However, it is time-consuming and challenging to verify whether a specific pending test is truly the last thing a patient needs prior to discharge (Safavi et al., 2022). Thus, the resulting lack of central transparency in patients’ discharge readiness can lead to the suboptimal allocation of resources that are essential to patient flow and the discharge of patients. These problems are further compounded by scale—a large hospital system typically has to manage hundreds of patient admissions and discharges each day. Although many hospital systems are investing in new technological or personnel strategies to address capacity challenges, such as “capacity physicians” (Safavi et al., 2022) or capacity command centers (Åhlin et al., 2023; Franklin et al., 2022; Franklin et al., 2023), the challenge of reliably identifying high-yield interventions remains.

To address this challenge in a systematic, data-driven way, we build upon the work described in Safavi et al. (2019) which introduced the terminology of *barriers to discharge* to dynamically represent the list of issues (e.g., “abnormal lab results”, “issues with mobility”, or “patient awaiting placement at a facility”) that could potentially stand in the way of discharging a patient. This framework uses clinical and administrative barriers as building blocks to construct a neural network model that can accurately predict how likely a patient is to be discharged within the next 24 hours. It outputs, for each patient, the likelihood of discharge together with a list of open barriers at the moment of prediction. This is important because the concept of barriers is central to how care teams approach the day-to-day management of inpatients, making the model’s output easily interpretable and actionable.

However, the model does not give any guidance on which of these barriers should be prioritized for those patients who are closer to discharge instead only highlights barriers which, if resolved, increase discharge likelihood. Moreover, retrospective analysis shows that not all barriers of a patient need to be resolved before discharge. Conceptually, this suggests that there exists a subset of *minimal barriers* whose collective resolution constitutes the minimal effort to enable a patient’s discharge. In this work, we combine machine learning models with a practically tractable integer programming approach to identify patients’ minimal barriers. This methodology uncovers targeted, data-driven interventions which may be the most effective to expedite patient flow by balancing the likelihood that barriers will be resolved with the possible change in discharge likelihood associated with their collective resolution.

### Approach—Discharge Prediction, Resolution Likelihood, and Minimal Barriers

Our approach is two-fold. First, we adapt the data representation of Safavi et al. (2019) in a way that allows us to incorporate sequential and logic-based dependencies among barriers in our prescriptive optimization model. We also train another set of machine learning models that predict, for each patient and for each of their open barriers, the likelihood it will be resolved prior to discharge. This step is critical given the complexity and variety of barriers. For example, some barriers are clinical in nature (e.g., waiting for patient to discontinue intravenous narcotics) and others resource-focused (e.g., waiting for post-hospital facility placement). Notably, even for a given barrier, there is heterogeneity across patients in whether the barrier will (or must) be resolved prior to discharge (e.g., some patients will go home on an oxygen support device while others will not). Secondly, these predictive models are combined within a mixed integer optimization model to answer the following question: among a patient’s numerous barriers, does there exist a small subset of those barriers which balances the likelihood that they are resolved prior to discharge *and* the likelihood of discharge for that patient given that those barriers are resolved? We call this the *minimal barriers* (MIN) identification problem. Intuitively, among a patient’s currently open barriers, the MIN should be defined as those which are both necessary and sufficient for discharge, i.e., they are likely to be resolved prior to discharge, and a patient’s discharge is likely to occur given that these barriers are resolved. We hypothesize that patients with discharge scores not too low nor too high (whom we formally define as “maybe” in Section 2.2) will account for most patients with non-empty MIN sets. We also consider how uncertainty in model predictions affects prescriptions, and we study a robust analogue of the nominal problem.

The central modeling challenge posed by such a problem is that this is not merely about prediction, as the outcome (a patient’s discharge) is dependent on actions (the resolution of barriers) which may or may not take place. Understanding and addressing this question is a critical step in achieving the broader aim of developing a transparent, centralized, scalable, and effective discharge process for hospitalized patients. Although our framework is more broadly applicable, in this work we limit our focus to surgical inpatients, i.e., patients who are cared for by a surgical service and require hospitalization following their surgical procedure.

#### 1.1 Contributions

Our main contributions can be summarized as follows:

1. We combine a discharge prediction model and barrier resolution likelihood models in a prescriptive mixed integer optimization model that identifies which barriers of a patient, if resolved, are most likely to increase discharge readiness. By jointly optimizing for both discharge likelihood and barrier resolution likelihood, we are able to select a small set of realistic and impactful barriers. We also develop a scalable iterative approach to solve this problem that finds optimal solutions for each patient in less than a second on average.
2. We present a detailed empirical assessment using real data to illustrate the effectiveness of the approach in identifying barriers for surgical inpatients at a large academic medical center. In doing so, we discuss how to calibrate our approach to identify a practically reasonable number of patients and barriers and show that our personalized barrier resolution likelihood approach is superior to a naïve approach that does not incorporate such information.
3. Finally, we study how estimation error in the discharge prediction model (namely, variance in prediction estimates) can contribute to changes in the minimal barrier set. We propose a robust optimization model which can be used to overcome these challenges, and demonstrate an application of our approach to the specific problem of identifying patients to prioritize for post-hospital facility placement.

### Implications for Practice

This study has several practical implications for hospital capacity management. As hospitals increasingly focus on creating centralized management to direct patient flow (such as capacity command centers), the modeling and algorithmic framework and tools we have created significantly enhance the ability to prioritize patients and specific actions per patient:

1. *Identify a small subset of barriers*: by incorporating the likelihoods that a patient’s barriers will be resolved prior to discharge, we are able to identify a small subset of barriers that both require resolution and for which resolution is likely to result in discharge. We show that our personalization approach is advantageous (relative to an appropriate baseline), in contrast to current practice which often uses a one-size-fits-all approach and **frequently ignores barrier resolution likelihoods entirely**. Importantly, we show that our approach can provide nontrivial insights about which barriers are frequent versus those which are identified in the minimal barrier set. This is especially important as certain high frequency barriers (such as imaging, which is often identified as a barrier) are rarely prescribed.
2. *Enable targeted interventions*: this prescriptive framework allows hospitals to pursue targeted, barrier-specific interventions. We demonstrate this for the use case of post-hospital facility (skilled nursing facility, rehabilitation hospital, or long-term acute care) placement. Even though many hospitals are investing in partnerships with post-acute facilities (or creation of their own facilities), they often lack insight into the operational workflow needed to identify which patients (across an entire hospital) should be prioritized for placement. Our approach addresses this challenge directly, and we estimate the potential benefit of our approach. While our proposal requires prospective clinical validation, we believe it forms a useful basis for numerous targeted interventions which can address the worsening hospital congestion that capacity management teams face.

#### 1.2 Related Literature

Our research is closely related to existing research in healthcare operations, interpretable machine learning, causal inference (particularly treatment personalization), and combinatorial multi-armed bandits. We briefly summarize the connections to such work and how our work differs.

### Healthcare operations

Inpatient capacity management and patient flow planning has been studied from a variety of perspectives. Conforti et al. (2011) and Gartner and Kolisch (2014) look at the problem from a hospital resource allocation perspective and employ mathematical models to maximize patient flow and contribution margins, respectively, while ensuring each patient goes through their respective clinical pathway. From inpatient bed capacity management perspective, some studies such as Kim et al. (2014) and McCoy et al. (2018) focus on predicting total discharge volume on a given day while others such as Liu et al. (2010), Azari et al. (2012), and Morton et al. (2014) focus on predicting length-of-stay (LOS) for each individual patient. There has also been work to understand discharge readiness as it progresses through a patient’s hospitalization, both manually (De Grood et al., 2016) and using machine learning (Pahlevani et al., 2024). More recently, Na et al. (2023) use machine learning to predict multiple patient outcomes including 24-hour and 48-hour discharge likelihoods. After deploying a webtool that communicates the patient outcome predictions and integrating it into the daily patient review processes at the study hospitals, they report a significant reduction in the average LOS of patients, which highlights the practical potential of such ML-driven models when implemented properly. In the present study, we extend our previous work (Safavi et al., 2019) in which we developed a neural network model to predict discharge likelihoods for surgical inpatients. We use this underlying model as one input in our novel prescriptive approach while also incorporating additional information about how likely a specific barrier is to be resolved by the time of discharge.

### Interpretable machine learning

Machine learning (ML) algorithms, able to process substantial electronic clinical and operational data, offer an opportunity to create systematic, scalable, and accurate predictions of which patients will be discharged. These algorithms, however, must be combined with an interpretable clinical representation to create an output that helps guide the clinical discharge processes. Rajkomar et al. (2018) proposed an automated patient representation which EHRs and construct relevant features in an unsupervised way. They demonstrate that deep learning methods using this representation are capable of accurately predicting multiple medical events such as readmission, prolonged length of stay from multiple centers without site-specific data harmonization. In contrast, Bertsimas et al. (2021) construct an expertise-driven patient representation on top of EHR data and apply an optimal classification tree machine learning approach to predict several aspects of patient flow such as mortality and discharge within 24 hours; they argue that the black-box nature of deep learning models impedes adoption from doctors and caregivers who are not engaged in the modeling process.

There has been a variety of work to explain black-box models (e.g., Ribeiro et al., 2016). However, these frameworks typically account only for which features contribute to the current prediction score, and as such, they do not answer the decision-making problem of “what interventions should be made (i.e., barriers resolved) such that a desired outcome is achieved?” Fischetti and Jo (2018) and Anderson et al. (2020) tackle this problem by answering the question “what is the minimal subset of features that needs to be modified in order to change the currently predicted class?” They present MIP formulations for high-dimensional piecewise linear functions that correspond to trained neural networks. Anderson et al. (2020) demonstrate the efficacy of their approach in a well-known benchmark image classification problem. From a methodological point of view, their work is the closest to the framework we present; several modifications (described later) have to be made to their approach to incorporate heterogeneity in barrier resolution likelihood.

### Personalized Treatments

In the causal inference literature, the *Potential Outcomes Framework* (Rubin, 2005) is often used to quantify the causal effect of a treatment for a given patient by comparing the potential outcomes. The standard framework was developed for a binary treatment (i.e., treatment or control), and has been adapted for multiple discrete or continuous treatments since. Recent approaches such as Shalit et al. (2017) and Bica et al. (2020) use machine learning to estimate personalized potential outcome function from historical observational data containing both contextual features and a treatment feature. In our setting, resolution of an open barrier could be thought of as a “treatment.” A major challenge with directly adapting such techniques here is that there are typically multiple (potentially tens of) open barriers for a given patient and more than one of them could require resolution prior to discharge (note, though, that there are some recent literature on multi-arm personalized treatment problems, e.g., Bertsimas et al., 2016). **Combinatorial Multi-armed Bandits:** Lastly, the class of *combinatorial multi-armed bandit* problems (CMAB) from reinforcement and online learning problems (W. Chen et al., 2016) share some similarities with our approach. In CMAB, the objective in each round is to choose a subset of base arms (collectively called a *super arm*) in a way that maximizes the expected gain. It is assumed that each arm’s properties are only partially known and may become better understood in the subsequent rounds only by choosing that arm. A variant of the problem is called Contextual CMAB (CC-MAB) which incorporates side information associated with each arm in each round (L. Chen et al., 2018). The exploitation phase resembles our problem in that it allows personalization (via context features) and combinatorial selection of multiple arms; however, they assume that the reward function is a known submodular function whereas in our case this must be estimated.

#### 1.3 Paper Structure

The remainder of the paper is structured as follows. Section 2 focuses on the modeling approach including study setting (2.1), discharge prediction model (2.2), barrier resolution models (2.3), and the prescriptive optimization model (2.4). Section 3 includes detailed empirical results when the prescriptive approach is applied to data from a large academic medical center. Section 4 focuses on the impact of uncertainty on model results and describes a model extension to address these shortcomings.

## 2 Modeling Approach

In this section, we describe the foundational elements of our modeling approach and how these are incorporated into a prescriptive model. We first describe the study setting and the data set that we use to test our models. We next describe how we change the data representation of the discharge prediction model (Safavi et al., 2019) to construct the barrier-resolution predictive models. We conclude by defining the optimization model that is used to derive the MIN barriers.

### 2.1 Study Setting

We use data from a 1019-bed academic medical center in the northeastern United States where more than 36,000 surgical procedures are performed annually. We include all adult (≥ 18 years old) inpatients who had a surgical procedure completed during their hospitalization, were cared for by a surgical team on an inpatient floor, and were discharged between May 1, 2016, to September 30, 2019. During the study period, there were 52,724 unique hospitalization episodes with average length of stay of 4.45 days. To mimic how a prescriptive model might work in practice, we capture, for a given patient on a given day, all data collected up until 23:59 on the day prior. This time was used because the clinical and administrative database is updated every 24 hours shortly after midnight. Our approach reflects a common limitation of real-world implementation, namely, that daily updates are the most common and feasible in practice. Therefore, we do not evaluate more frequent data nor model updates.

Our central unit of observation is a patient-day pair, and our outcome of interest is whether the patient was discharged within 24 hours (i.e., on that calendar day). During the study period, the the model made predictions on an average of 188.2 patients per day, amounting to overall 234,649 patient-day pairs. The average number of discharges per day was 42.3. We temporally split the data intro three sets: training, validation, and test. The training set includes all observations of patients admitted on or before December 31, 2018 (78.9% of patients and 78.2% of patient-days); the validation set includes patients admitted between January 1, 2019 and April 30, 2019 (9.5% of patients and 10.0% of patient-days); and the test set includes admissions during May 1, 2019 and September 30, 2019 (11.6% of patients and 11.8% of patient-days). All results presented are based on test set (out-of-sample) performance using models built from the training and validation data. This study was conducted with appropriate approval from the Mass General Brigham Institutional Review Board (protocol 2011P001124).

### 2.2 Discharge Prediction Model and Data Representation

In earlier work, we trained and clinically validated machine learning models to reliably predict whether a given patient will be discharged from the hospital within the next 24 hours (Safavi et al., 2019), ultimately selecting a simple feed-forward neural network (with a single hidden layer with 100 nodes) which outperformed others across all evaluation metrics. This approach takes as its input a variety of clinical and administrative data extracted from a hospital’s EHR and provides a prediction score (in [0, 1]) as its output which can be interpreted as the likelihood that the patient will be discharged within 24 hours. Input data included both *static* features invariant during the hospitalization (e.g., patient age) and *dynamic* features like laboratory values, medications, and orders (Table SM1); they were designed and selected based on whether the clinical care teams currently use it to evaluate a patient’s discharge readiness, the reliability of the data, and the consistency of usage by different clinical services.

Central to this approach is the concept of **barriers**, which are defined as any event that may prevent a patient from being discharged from the hospital. A barrier could be specific to a type of surgical procedure or generalizable across multiple types of surgical procedures; a total of 149 potential barriers were identified in Safavi et al. (2019) by a multidisciplinary team that included physicians, nurses, case managers, physical therapists, and nutritionists. We use the same underlying data, but we modify the feature representation in a way that is amenable to a prescriptive optimization framework. In particular, we represent the dynamic status of a patient’s barriers by using a *trigger-resolution* framework where each barrier is encoded by a *pair* of binary features: (1) a trigger feature that marks whether the barrier has been activated during the hospitalization; and (2) a resolution feature that signals whether the barrier has been resolved. Barriers that have been triggered but are not yet resolved are called **open**. Feature vectors for observation *i* are represented as **x**_*i*_ = (**s**_*i*_, *d*_*i*_, **t**_*i*_, **r**_*i*_), composed of four components: static, days since admission, trigger, and resolution features, respectively. For the present study, we retrained the discharge prediction model using this modified representation; performance was similar (see Table SM2).

#### Calibration

To ensure appropriate model interpretation of predictions as probability estimates, we must verify that the model is properly *calibrated* (Steyerberg, 2019). While there are numerous ways to assess model calibration, we use Brier scores and the Spiegelhalter *Z*-test (Huang et al., 2020). To achieve calibration, we applied Platt scaling to the trained discharge prediction model. For full details and calibration plots, see SM1.3.

#### Yes/Maybe/No framework

To understand how our prescriptive model performs, we stratify performance by a patient’s baseline discharge score into three groups: patients who are likely to be discharged (Yes patients); patients who are unlikely to be discharged (No patients); and patients whose discharge score is neither high nor low (Maybe patients). This approach better reflects clinical and operational ambiguity in discharge predictions beyond the classical binary dichotomization. Formally, we define these groups for a discharge score *s* as No for *s* ∈ [0, *S*_−_), Maybe for *s* ∈ [*S*_−_, *S*_+_], and Yes for *s* ∈ (*S*_+_, 1], where *S*_−_ and *S*_+_ are fixed thresholds defined so that the No and Yes categories have high negative and positive predictive power (NPV/PPV), respectively. Precisely, *S*_−_ was chosen so that 95% of the (training sample) predictions that fall under No category were not discharged within 24 hours (NPV=95%); and *S*_+_ was chosen so that 75% of the predictions that fall under Yes category were discharged within 24 hours (PPV=75%). These percentages were based on clinical input about tradeoffs of making the respective predictions. Based on the training and validation data, we set the thresholds as *S*_−_ = 0.19 and *S*_+_ = 0.59. The performance of these choices on the test data is shown in Table 1.

**Table 1:** Discharge prediction categories and comparison of actual discharge times.

| Prediction category | # (%) of Observations | Leave within 24 hours | Leave in 24-48 hours | Leave after 48 hours |
| --- | --- | --- | --- | --- |
| NO | 16373 (59.3%) | 823 (5.0%) | 2017 (12.3%) | 13533 (82.7%) |
| MAYBE | 7980 (28.9%) | 2933 (36.8%) | 2167 (27.1%) | 2880 (36.1%) |
| YES | 3248 (11.8%) | 2367 (72.9%) | 548 (16.9%) | 333 (10.2%) |

Note that the Yes and No categories make up 71.1% of all predictions. Moreover, these two correspond informally to “leave within 24 hours” and “leave after 48 hours,” respectively. On the other hand, most of the patients who leave between 24-48 hours fall under the Maybe category. Combining this with the insight that “false positive patients without a clinical reason to stay are typically discharged within 24 to 48 hours” (Safavi et al., 2019), the Maybe category roughly corresponds to the potentially actionable patients for the purposes of saving bed-days. Therefore, we hypothesized that the Maybe patients would be the most prevalent patients to have specific MIN identified.

### 2.3 Barrier Resolution Prediction Models

We use 149 clinical and operational barriers (see Table 2 for 10 example barriers). Note the significant heterogeneity in barrier occurrences and resolutions; some barriers such as “pending physical therapy (PT) discharge disposition recommendation,” “irregular diet,” and “on intravenous narcotics” occur in many patients, while others mostly occur only in a smaller fraction (for example, only 1.3% of patients have an “on heparin” barrier during their hospitalization). There is also heterogeneity in the percentage of patients for whom the respective barrier is resolved prior to discharge. The data in Table 2 imply that not all barriers are equally likely to be resolved prior to discharge. Moreover, the importance of a given barrier’s resolution for a patient’s discharge is not identical across different patients, specifically, for some the barrier might be critical to allow discharge, whereas for others it might not prevent the discharge (in the example earlier, a patient who is on home oxygen support prior to admission can likely be discharged on home oxygen support). Overall, only 43.6% of the barriers which get triggered at some point during the hospitalization of the patient get resolved by the time of discharge.

**Table 2:** Ten barriers and their respective out-of-sample summary statistics. “Patient level” indicates the percentage of patients for whom the barrier was triggered (and subsequently resolved prior to discharge), while “Patient-day level” provides the percentage of hospital days for which the barrier was activated and then the percentage where it was resolved (after activated).

| Barrier | Patient level |  | Patient-day level |  | Resolution model |  |
| --- | --- | --- | --- | --- | --- | --- |
|  | % triggered | % resolved | % open | % resolved | AUC | Brier score |
| Pending Physical Therapy’s discharge recommendation | 96.0 | 46.3 | 59.5 | 45.4 | 0.91 | 0.12 |
| On oral narcotics | 69.2 | 24.9 | 49.7 | 21.2 | 0.72 | 0.15 |
| Irregular diet | 58.8 | 58.2 | 44.4 | 40.6 | 0.87 | 0.14 |
| Urinary catheter | 42.7 | 84.9 | 25.8 | 78.4 | 0.79 | 0.13 |
| On intravenous narcotics | 37.2 | 83.8 | 13.5 | 85.9 | 0.85 | 0.10 |
| On oxygen device | 34.8 | 93.6 | 22.4 | 83.7 | 0.77 | 0.12 |
| Low urine output | 34.7 | 87.0 | 17.1 | 83.9 | 0.89 | 0.08 |
| X-ray study | 33.9 | 80.6 | 20.7 | 75.3 | 0.70 | 0.17 |
| Awaiting post-hospital facility acceptance | 15.6 | 78.8 | 18.5 | 82.2 | 0.80 | 0.11 |
| On heparin | 1.3 | 94.9 | 0.5 | 92.5 | 0.58 | 0.08 |

To capture this heterogeneity, we build a predictive model for every barrier to predict the likelihood that it will be resolved prior to discharge given a patient’s current data. To do so, we use the same feature vector representation as above and define a binary outcome of whether the barrier is resolved prior to patient discharge. For each barrier, we include as training data the subset of (training) patients for whom that barrier is open. Our objective in creating such models is to balance model discriminative performance (i.e., high AUC) while ensuring that appropriate calibration on the training and validation sets. To accomplish this, we use (regularized) logistic regression; see SM1.4 for details and discussion of the relationship between the number of observations with a given barrier and out-of-sample AUC.

As a result, for each observation, we obtain the personalized barrier resolution likelihood vector, denoted **L** ∈ [0, 1]^149^, by concatenating the prediction scores of all resolution prediction models (*L*_*j*_ represents the resolution likelihood for barrier *j*; the dependence on the observation is omitted). In initial model-building, we also evaluated a multi-output neural network model (to predict joint resolution of all 149 barriers), but this approach performed significantly worse than simple models on the single-barrier-resolution prediction task. Therefore, we restrict our attention to the barrier-by-barrier approach described above.

### 2.4 Prescriptive Model—Identifying Minimal Barriers

We now describe how these predictive models can be combined to formulate and solve the MIN problem. As noted above, not all barriers of a patient are resolved prior to discharge. Moreover, among the barriers that are resolved before discharge, there may be some whose resolutions were not necessary for discharge. Conceptually, this suggests that there exists a subset of barriers (we call them *minimal barriers*, or MIN) whose collective resolution constitutes the minimal effort to enable a patient’s discharge. Our goal is to identify such a set of barriers.

To define our objective, we start with an idealized model. In particular, for a subset *S* of a patient’s current barriers, let ℙ_res_(*S*) denote the (idealized) probability that all the barriers in *S* are resolved prior to discharge, and let ℙ_dc_(*S*) denote the (idealized) probability that the patient is discharged within 24 hours *given* that the *S* barriers are resolved. We aim to solve the problem of finding *S* that maximizes

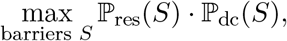

the product of the probability of the barriers being resolved and the probability of discharge given that resolution. An optimal barrier set *S*^*^ to this problem balances the likelihood of those barriers being resolved with the contribution that such barrier resolution makes to a patient’s discharge likelihood. The advantage of such an objective is that it implicitly accounts for the fact that some barriers are unlikely to be resolved even if they contribute significantly to discharge likelihood, while other barriers might be likely to be resolved but make little or no difference to a patient’s progression towards discharge. To solve this idealized problem, we need a way of estimating the two central quantities ℙ_res_ and ℙ_dc_. The former can be estimated using the barrier resolution prediction models described in Section 2.3 and the latter can be estimated using the discharge prediction model described in Section 2.2.

We now translate this idealized problem of identifying a patient’s *minimal barriers* into an optimization problem where the decisions on the open barriers (“whether the barrier should be selected to resolve”) are reflected as changes in the current feature vector **x**^*c*^ of a patient to a modified vector **x** (in this section, we suppress the index for observation *i* to make the exposition simpler). Consequently, we try to decide which features to modify, by allowable changes, so that the product of the following two objective functions is maximized:

- *J*_*r*_(**x**; **x**^*c*^): the product (among the selected barriers *S* represented in changing **x**^*c*^ to **x**) of each of the likelihoods that the given barrier will be resolved before discharge; this approximates ℙ_res_(*S*), the probability of the barriers in *S* being resolved, assuming all barriers resolve independently of one another. (We return to the independence assumption later.)
- *J*_*d*_(**x**; **x**^*c*^): the discharge likelihood within 24 hours if all selected barriers are resolved; this approximates the idealized ℙ_dc_.

Using these two objectives, we define the composite objective function *J* as the product *J* = *J*_*r*_ *· J*_*d*_ and we call this the **overall likelihood given a subset of barriers**. This is directly analogous to the idealized objective, with *J*_*r*_ estimating ℙ_res_ and *J*_*d*_ estimating ℙ_dc_. Instead of optimizing *J* directly, we adopt a constraint-based multiobjective optimization perspective. In particular, we compute the Pareto frontier for *J*_*r*_ and *J*_*d*_ by solving instances of maximizing *J*_*r*_ subject to (iteratively updated) lower bounds on *J*_*d*_. We proceed by describing the structure of the two different function *J*_*r*_ and *J*_*d*_.

#### Structure of *J*_*r*_

Notice that the first objective function *J*_*r*_ can be calculated by multiplying the personalized resolution likelihoods **L** of the selected barriers readily available from the barrier resolution prediction models described in Section 2.3. To linearize *J*_*r*_, we take the natural logarithm of the personalized likelihoods log(*L*_*j*_) so that maximizing the sum of log-likelihoods of the selected barriers would be equivalent to maximizing the likelihood that all selected barriers will be resolved, which we refer to as their *collective resolution likelihood*. Addition of a new barrier to the list of selected barriers always reduces the collective resolution likelihood. Therefore, this objective function implicitly favors reducing the number of selected barriers (i.e., parsimony).

The advantage over simply minimizing the number of selected barriers is that this way we are able to account for the differences in the resolution likelihoods of barriers. It is intuitive to conclude that one would prefer selecting a list of two barriers both very likely to be resolved by the time of discharge rather than a single barrier which will almost surely not be resolved. In this way, *J*_*r*_ can be generally thought of as a measure of **cost** (higher *J*_*r*_ corresponding to lower cost), and in other settings we expect that *J*_*r*_ might be more directly defined using actual costs.

#### Structure of *J*_*d*_

On the other hand, optimizing with the second objective function, *J*_*d*_, is a more challenging task for two reasons: it is implicitly expressed through a trained neural network; and the underlying decision space is discrete (i.e., binary decisions for whether each barrier is resolved). To address this, we use the work of Anderson et al. (2020) who proposed and evaluated tractable mixed integer program formulations to find the minimal subsets of features that leads to a desired change in neural network predictions. To apply their framework, we address two specific modifications: not all features are modifiable (only the resolution features of open barriers can be changed from 0 to 1); and the objective is not to minimize the number of selected barriers but instead to maximize the function log(*J*_*r*_) subject to the prediction score being at least some threshold *z*_0_. In SM2, we include a detailed exposition showing how these modifications can be made to yield a linear MIP formulation of the subproblem

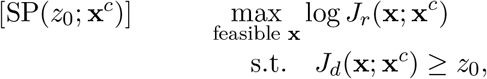

where *z*_0_ ∈ [0, 1] is a fixed threshold and **x**^*c*^ is a patient’s current feature vector. (The underlying decision variables are binary and capture whether each barrier is resolved.) Note that the key to this reformulation is using the underlying neural network structure of the model for predicting discharges. The resulting formulation has 2*h* + *p*_*b*_ variables and 4*h* + 2*p*_*b*_ + 1 constraints, where *h* is the dimension of the hidden layer in the neural network (in our case, 100) and *p*_*b*_ is the number of barriers (in our case, 149).

#### An Iterative Algorithm to Optimize *J*

Given the formulation of SP as a MIP which is amenable to commercial solvers, we are now ready to proceed with an algorithm for maximizing *J* for a given observation **x**^*c*^. As noted earlier, we do so by solving SP(*z*_0_; **x**^*c*^) across a spectrum of thresholds *z*_0_ ∈ [0, 1]. The corresponding algorithm is shown in pseudocode in Algorithm 1, which we refer to as the *Barriers-to-Discharge* (or BtD) algorithm. The optimal solution **x**^*^ to a given feature vector **x**^*c*^ identifies a set of **prescribed (or minimal) barriers**, namely, the MIN set. The algorithm follows the usual approach used when computing Pareto frontiers, namely, instead of solving SP(*z*_0_; **x**^*c*^) for all choices of *z*_0_ ∈ *{*0, *c*, 2*c*, …, 1*}*, we only solve it for a (typically smaller) set of values. In the remainder of the paper, we make use of two important definitions:

##### Algorithm 1

BtD(x^*c*^, l, *c*)

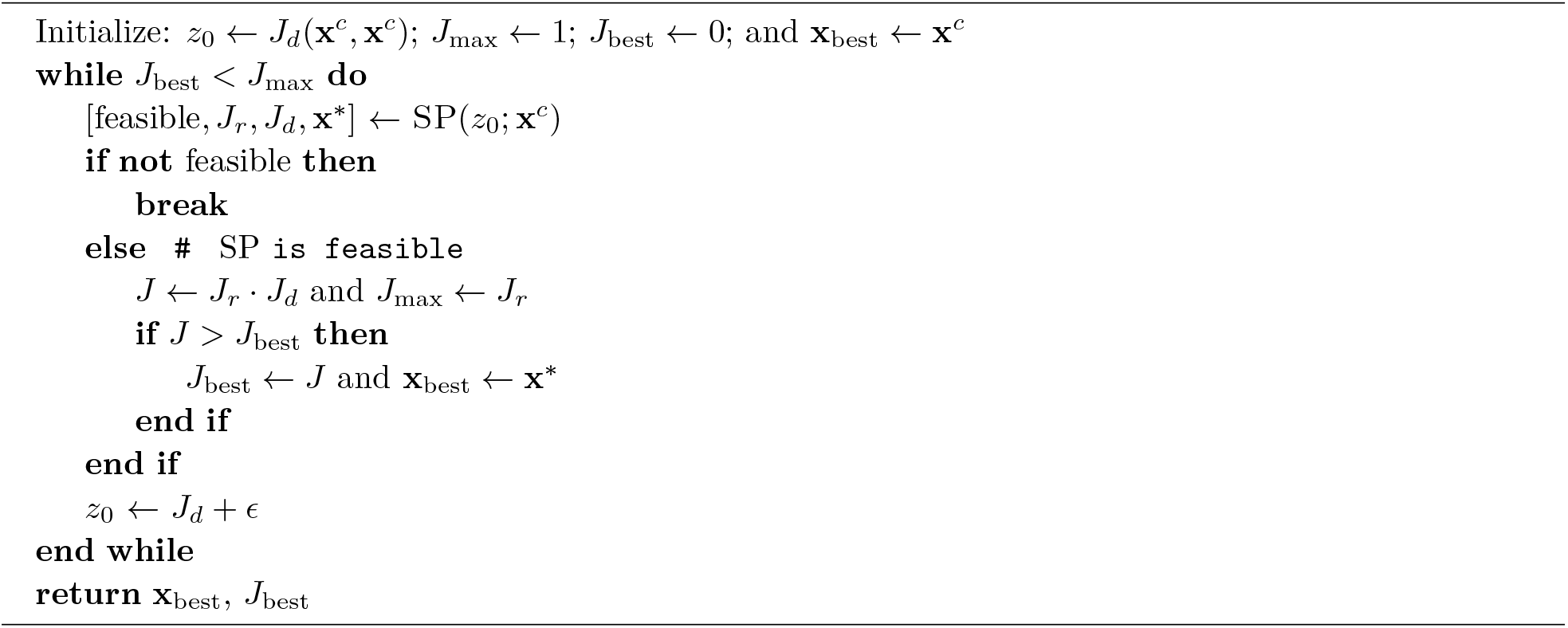

##### Definition 1

(Feasible patient). *Given a patient observation with corresponding feature vector* **x**^*c*^, *we say that the observation is* ***feasible*** *if there exists some set of open barriers such that J is strictly improved (up to tolerance c) relative to the value of J when no open barriers are resolved; an infeasible patient has no prescribed barriers. In mathematical notation, this is equivalent to there existing some feasible feature vector* **x** *such that J* (**x**; **x**^*c*^) *> J* (**x**^*c*^; **x**^*c*^) = *J*_*d*_(**x**^*c*^).

##### Definition 2

(Actionable patient). *Given a feasible patient with feature vector* **x**^*c*^, *we say that the patient is* ***actionable with likelihood*** *J* ^*^, *where J* ^*^ := *J* (**x**^*^; **x**^*c*^) *as computed via the BtD algorithm. For every J* ^*I*^ *< J* ^*^, *we say that the patient is* ***actionable above threshold*** *J* ^*I*^.

## 3 Prescriptive Model Evaluation

In this section, we assess the performance of our proposed approach and answer the following questions:

1. How does the composite objective function *J* perform in discriminating likelihood of discharge given a selected subset of barriers?
2. Does the BtD algorithm primarily identify Maybe patients as those who are actionable above certain thresholds as initially hypothesized? How does a patient’s actionability relate to their initial discharge score, and how many barriers are typically prescribed?
3. What lower threshold *J*_min_ should be chosen for actionable patients to yield a practically meaningful number of patients on a daily basis? Which barriers tend to be prescribed as minimal barriers, and how should these be interpreted in this context?
4. What is the contribution of personalized resolution likelihoods?
5. How does the BtD approach compare to complete enumeration in terms of computational efficiency? How can our approach be modified in settings with other predictive models?

We conducted our analysis on the out-of-sample test sample, i.e., 27,601 observations (patient-days) from 6,123 unique patient hospitalizations. We implemented the BtD algorithm in julia 1.10.4 and utilized Gurobi 11 as a mixed integer optimizer, with default parameters and a single thread, to solve the BtD algorithm. All experiments in this section were carried out on a laptop with processor 3.1 GHz Dual-Core Intel Core i5, memory 8 GB 2133 MHz LPDDR3 (experiments in Section 4 were run on a cluster computing environment to accommodate large-scale replications).

### 3.1 Efficacy of the Composite Objective Function

We begin by assessing how well the composite objective function *J* performs in capturing the idealized definition of MIN. Such idealized barriers should satisfy two conditions simultaneously: (*I*1) that they all must be resolved by the time of discharge and (*I*2) the patient must be discharged on the same calendar day that all of them are resolved. To test this, we performed the following analysis: for each test set observation, we randomly selected a subset of up to five of the patient’s open barriers and calculated the objective function value *J* corresponding to the modified feature vector obtained by resolving the selected subset of barriers; then, retrospectively, we labeled each instance as follows:

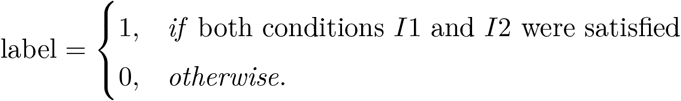

In this experiment 17.1% of observations met *I*1, 20.1% met *I*2 given that *I*1 was met, and 3.4% met both *I*1 and *I*2 (i.e., had a label of 1). We calculated the discriminative performance of *J* in terms of AUC for this labeled outcome. Overall, the AUC was 0.854 (95% CI [0.843,0.864]); within the Yes/Maybe/No subgroups, Yes patients had an AUC of 0.938 [0.924,0.952], Maybe patients 0.910 [0.899,0.922], and No patients 0.814 [0.799,0.829]. This suggests that *J* is particularly effective at discriminating between patients’ labels within the Yes and Maybe subsets of patients. (See SM1.6 for a discussion of *J* as a function of the number of selected barriers.)

### 3.2 Identifying actionable patients

To better evaluate the BtD algorithm and assess our hypothesis that Maybe patients account for the majority of actionable patients, we solved the BtD algorithm for all 27,601 observations. The corresponding results regarding the percent of patients who are actionable at different likelihood thresholds *J*_min_ ∈ *{*0, 0.25, 0.5, 0.75*}* are shown in Figure 1.

**Figure 1:**
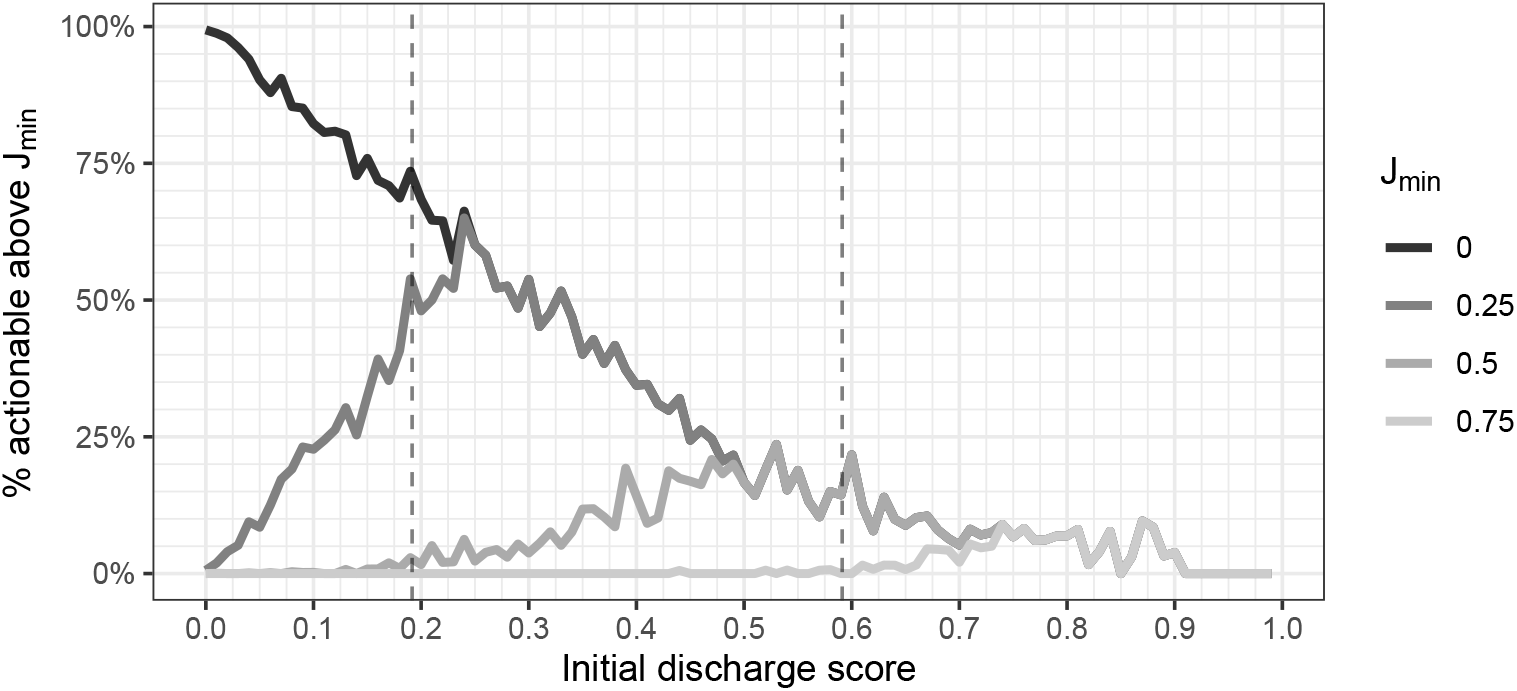
Percentage of actionable patients at a given score and with given value of *J*_min_, where *J*_min_ ∈{0, 0.25, 0.50, 0.75}. Two vertical lines for Maybe patient score cutoffs are shown; note that *J*_min_ = 0 corresponds to feasible patients (initial scores grouped to second decimal).

#### Feasibility

The percentage of patients at or above a given discharge score who are *feasible* is shown with *J*_min_ = 0 in Figure 1. We observe that the percentage of feasible patients generally decreases linearly with increasing baseline discharge scores. As a given observation being feasible coincides with the existence of at least one barrier in MIN (per the BtD algorithm), this behavior is consistent with our expectations of how MIN would behave: for patients who are unlikely to be discharged in their current state, there likely exist some barriers which could raise their discharge likelihood (while also being reasonably likely to be resolved, given that the patient is further from discharge); at the same time, patients with increasingly higher scores are more likely to be discharged in their current state, so it is less likely that there exists barriers which not only increase the patient’s discharge likelihood, but importantly are also likely to be resolved. This is particularly important given the patient heterogeneity noted earlier.

#### Actionability at different thresholds

Not surprisingly, increasingly larger minimum thresholds *J*_min_ affect the percentage of patients who are actionable. We also observe that the percentage actionable above threshold *J*_min_ peaks at approximately an initial discharge score of *J*_min_ across the different choices shown. Overall, the qualitative behavior seen here suggests that the optimization approach we have taken to identifying actionable barriers aligns with the clinical intuition we used to approach this problem, namely, that the composite objective would capture the trade-off between the likelihood of improving a patient’s discharge and the likelihood of being able to resolve barriers to put the patient in that improved discharge state. We also note that the changing thresholds *J*_min_ highlight the inherent tradeoff between the number of patients identified as actionable above such a threshold and how likely patients are to be discharged with such barriers resolved. We return to this point in Section 3.3.

#### Proportion of Maybe patients

This analysis confirms our initial hypothesis: while Maybe patients account for 28.8% of observations in the test set (Table 1), they represent a disproportionately large share of actionable patients across a range of thresholds. For example, at a threshold of *J*_min_ = 0.25, Maybe patients account for 60.6% of actionable patients, while at a threshold of *J*_min_ = 0.5, Maybe patients account for 74.5% of actionable patients. Not surprisingly, though, we do see that for *J*_min_ sufficiently large (in our case, *J*_min_ ≥ 0.60), Maybe patients are no longer the majority of those identified as actionable (the same is true of *J*_min_ sufficiently small, in our case *J*_min_ ≤ 0.20).

#### Number of prescribed barriers

Among all feasible patients, the number of open barriers is 11.5 on average (median of 10, range of 1-40), while the number prescribed for such patients is 3.0 on average (median of 2, range 1-27). This suggests that the BtD algorithm is effective at reducing the list of barriers for each patient to a more reasonable number (26.9% of the list of open barriers on average, median 21.4%, range [3.4%,100%]). Consistent with this behavior, the number of prescribed barriers differs noticeably across the patient groups: No patients are prescribed an average of 3.5 barriers (median 3, range [1,27]), Maybe patients 1.2 barriers (1, [1,4]), and Yes patients 1.0 barriers (1, [1,2]). Finally, as the number of open barriers is negatively correlated with initial discharge score (correlation coefficient of −0.402 for feasible patients), we expected that the number of prescribed barriers would also be negatively correlated with initial discharge score; indeed, we found this to be the case (correlation coefficient of −0.462).

### 3.3 Model performance—Thresholds for actionability and barriers selected

Given an overall understanding of how actionability relates to a patient’s underlying discharge likelihood, we now focus on addressing some specific questions: what choice of *J*_min_ gives a reasonable number of actionable patients which is practically feasible to review on a daily basis; which barriers are typically prescribed (and how should these be interpreted); and how often is a single prescribed barrier the one with the highest personalized resolution likelihood?

#### Model performance across choices of *J* _min_

To assess the model performance, we consider several key metrics (aligning with those considered in Section 3.1):

(*M* 1) the percentage of actionable observations for whom selected barriers were all resolved by the time of discharge;

(*M* 2) the percentage of observations meeting criterion *M* 1 who were discharged on the same day that all selected barriers were resolved; and

(*M* 3) the percentage of observations for which both *M* 1 and *M* 2 were satisfied.

We evaluated these across various thresholds *J*_min_; the corresponding results are shown in Table 3. We also include several additional variables, such as values of the composite objective function and the percent of actionable patients relative to all patients.

**Table 3:** Prescriptive model’s test-set performance at different choices of *J*_min_. The percent shown next to the number actionable is relative to all test-set observations. Averages for 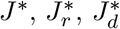, and “# Barriers” are based on those values as computed using the BtD algorithm.

| $J_{\min}$ | # Actionable<br>above $J_{\min}$ | | # Barriers | | Average value | | | Metric (%) | | |
| --- | --- | --- | --- | --- | --- | --- | --- | --- | --- | --- |
| | | | Open | Prescribed | $J_r^*$ | $J_d^*$ | $J^*$ | $M1$ | $M2$ | $M3$ |
| 0.0 | 18619 | (67.5%) | 11.50 | 3.02 | 0.67 | 0.25 | 0.18 | 68.7 | 33.2 | 22.8 |
| 0.1 | 10872 | (39.4%) | 9.42 | 2.00 | 0.75 | 0.36 | 0.28 | 74.1 | 35.9 | 26.6 |
| 0.2 | 6590 | (23.9%) | 8.85 | 1.67 | 0.80 | 0.45 | 0.37 | 76.8 | 40.5 | 31.1 |
| 0.3 | 3934 | (14.3%) | 8.36 | 1.45 | 0.84 | 0.53 | 0.45 | 78.9 | 45.6 | 35.9 |
| 0.4 | 2195 | (8.0%) | 7.85 | 1.33 | 0.87 | 0.61 | 0.53 | 79.6 | 49.3 | 39.3 |
| 0.5 | 1074 | (3.9%) | 7.33 | 1.22 | 0.90 | 0.68 | 0.61 | 79.7 | 54.3 | 43.3 |
| 0.6 | 483 | (1.7%) | 6.89 | 1.13 | 0.92 | 0.76 | 0.70 | 77.6 | 61.1 | 47.4 |
| 0.7 | 204 | (0.7%) | 6.98 | 1.08 | 0.94 | 0.82 | 0.77 | 78.9 | 72.7 | 57.4 |
| 0.8 | 64 | (0.2%) | 7.06 | 1.05 | 0.95 | 0.88 | 0.84 | 85.9 | 80.0 | 68.8 |

As expected, we see that the percent of actionable patients who have all prescribed barriers resolved **and** are discharged on the same day as their resolution (i.e., metric *M* 3) increases as the threshold for actionability *J*_min_ is increased, highlighting the tradeoff between the number of actionable patients identified and the percent of those patients for whom the prescribed barriers meet metric *M* 3. In our setting, with a median daily surgical patient census of 190 patients in the test period, we believe that a reasonably sized list of actionable patients would account for 3-5% of patients (approximately 5-10 patients); therefore, *J*_min_ = 0.5 is a reasonable choice.

Overall, 67.5% of all patients are feasible. We believe that infeasible patients are likely those at an ambiguous stage in their care path, with some of the barriers they will encounter during their hospitalization not even been triggered yet (e.g., placing a request for a post-hospital facility). The reason we used the *trigger-resolution* framework to represent barriers was that the introduction (trigger) of a barrier in itself will likely reduce the discharge prediction score, but it will also make possible a resolution whose contribution—if it occurs—to the prediction score will be higher than the score reduction caused by the triggering of the barrier. Indeed, if we compare feasible and infeasible patients, feasible patients tend to have more open barriers: overall an average of 11.5 barriers versus 7.0, difference of 4.5, 95% CI [4.4,4.6]. This difference is partly attributable to feasible patients having lower baseline discharge scores, therefore we also looked at Yes/Maybe/No patients separately as well: among No patients, feasible patients have 3.0 more barriers on average relative to infeasible patients (12.3 versus 9.3, 95% CI [2.8,3.3]); among Maybe patients, feasible patients have 1.1 more barriers on average (8.2 versus 7.1, 95% CI [1.0,1.3]); and among Yes patients, feasible patients have 1.4 more barriers on average (7.1 versus 5.7, 95% CI [1.1,1.6]).

To conclude the quantitative discussion of the model performance, it is important to compare the values of the objective functions and their respective metrics. Per Table 3, we have the following:

1. The value 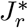 corresponds to a barrier resolution likelihood and is a proxy of metric *M* 1. We see that on average, 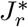 consistently overestimates *M* 1 across most thresholds. The cause of this is likely multifactorial in nature. Firstly, note that 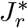 is defined as a product of individual likelihoods, while *M* 1 captures a **joint** resolution percent; however, barrier resolution is likely **not** independent across barriers, and therefore the product of individual estimates performs poorly. Interestingly, if we restrict our attention to actionable patients with a single prescribed barrier, the average value of 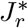 still overestimates *M* 1. This suggests that, even though the resolution likelihood models were calibrated on a barrier-by-barrier basis, the optimization model has induced a form of selection bias such that the selected barrier has an overly optimistic resolution estimate. Nevertheless, it is important to recall that Table 3 is only for *actionable* patients; therefore, patients with barriers with all low resolution likelihoods (and hence are infeasible) are not reflected here. Indeed, for lower values of *J*_min_, 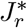 is a well-calibrated proxy for *M* 1.
2. The average score 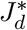,in contrast, underestimates the 24-hour discharge percentage *M* 2 for small values of *J*_min_ and overestimates *M* 2 for larger values of *J*_min_, and the latter discrepancy is to a larger degree than the one noted for *M* 1 (hence *M* 3 is also overestimated). In Section 4, we extend our model to better calibrate estimates of *M* 3 using a robust optimization approach, particularly for higher values of *J*_min_ that are of practical interest.

#### Barrier prescription behavior

To facilitate a qualitative understanding of the BtD algorithm, we also considered which barriers were prescribed by the model, especially in relation to the prevalence and average resolution likelihood of that barrier. The results of this analysis are shown in Figure 2 for the subset of patients who are actionable at *J*_min_ = 0.5 (the barriers in Table 2 are labeled for context). First of all, we note that barriers with low average resolution likelihood (below approximately 35%) are rarely prescribed in MIN. (An example of such a barrier is “PO Narcotics”, denoting that the patient is taking oral narcotic medication.) Further, with the exception of seven barriers, the model almost exclusively prescribes a lower percent of that barrier than the average resolution likelihood.

**Figure 2:**
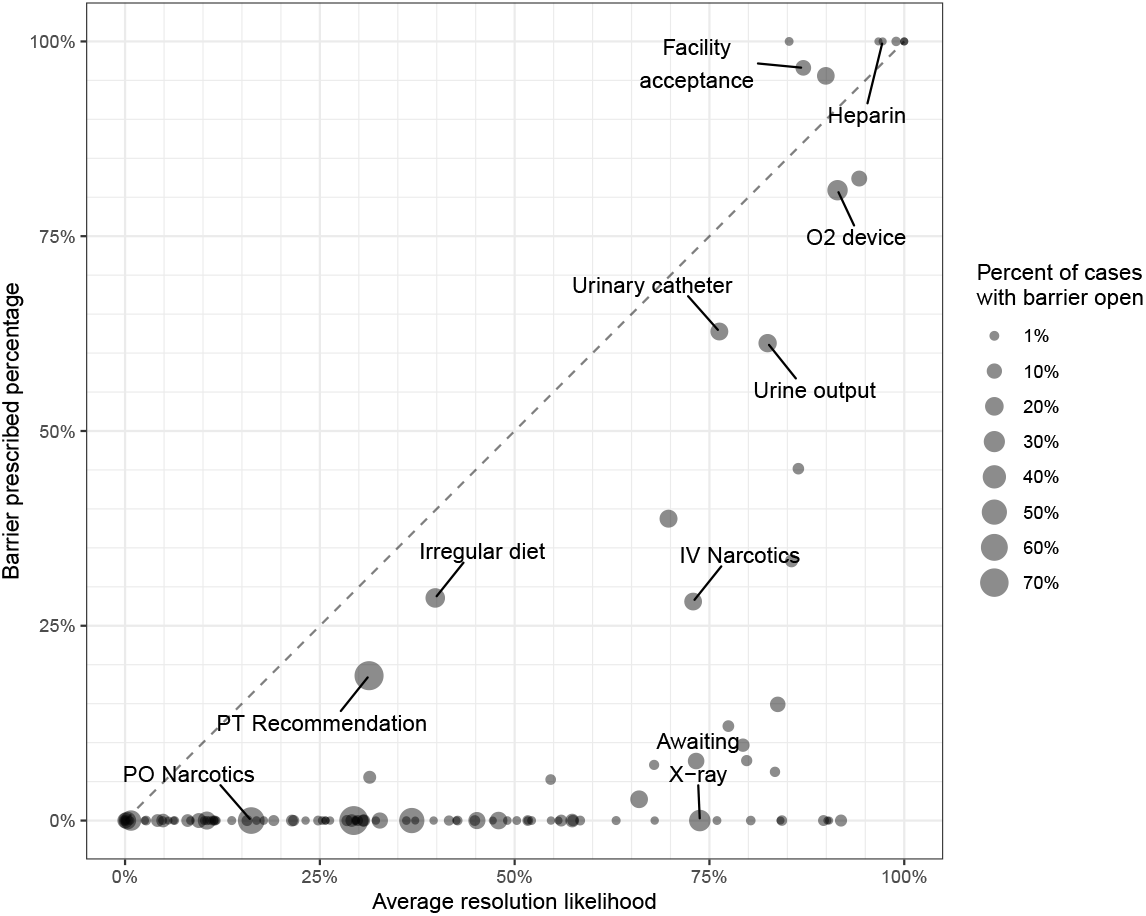
Average barrier resolution likelihood and prescription percentage among patients who are actionable above *J*_min_ = 0.5. Each point is a barrier and its size corresponds to the percent of patients with that barrier triggered. All percentages are relative to this population only (with resolution/prescription percents relative to the subset of those with such a barrier). Barriers which are never open in this population are not shown. Abbreviations: PO=“medication taken by mouth”; IV=“intravenous”; PT=“physical therapy”; O2=“oxygen.” confirms that imaging studies are often barriers, they are rarely prescribed (given low estimated resolution likelihoods), further highlighting the importance of incorporating resolution likelihood into the modeling framework.

This figure also highlights the relationship between some of the barriers, and it also points to the observational nature of the data. For example, let us consider two barriers related to post-discharge accommodations: “Facility acceptance” (indicating that the patient is awaiting acceptance in a post-hospital facility, such as a skilled nursing facility, to which they will be transferred upon discharge) and “PT Recommendation” (indicating that the patient is still awaiting the physical therapy team to provide a recommendation on the appropriate post-discharge location for the patient). Among actionable patients above *J*_min_ = 0.5, Facility Acceptance is the fifth most highly prescribed barrier among all 149, highlighting the critical importance of post-discharge facilities for a nontrivial number of patients (for those with the barrier, it is prescribed 96.6% of the time).

In contrast, in this same group the PT Recommendation barrier is prescribed 18.6% of the time it is open for a patient. We believe this discrepancy is likely due to the fact that the PT Recommendation barrier is not being reliably documented as resolved by discharge (as, in reality, the patient likely did receive such a team’s input before discharge), hence lower resolution likelihoods are estimated and, as a result, it is less likely to be prescribed. Overall, this highlights the inherent challenge with using EHR data, where documentation patterns and behavior evolve over time. Lastly, we believe that Figure 2 could also serve as the basis for further discussion with clinical teams on the ground to identify sources of the discrepancies between resolution versus prescription percentages. For example, imaging studies (such as magnetic resonance imaging, or MRI) are often identified by care teams as a barrier to patient progression (Safavi et al., 2022). While our data

Finally, we conclude this qualitative assessment by noting that we made the deliberate choice to include both clinical and non-clinical barriers in our model. Examples of clinical barriers include things like “O2 Device” (indicating a patient needs oxygen support), while “Facility Acceptance” or “Awaiting MRI [Magnetic Resonance Imaging]” are non-clinical, resource-focused barriers. This choice was made early in the model development stages, and the primary impetus for that was because, even if specific interventions might be targeted at non-clinical barriers (for example, by facilitating additional MRI resources on a given day), the context of what clinical barriers remain for a patient is critically important. For example, if a patient is prescribed both a clinical and a non-clinical barrier, and their non-clinical barrier is resolved but the clinical barrier remains, the patient is still not necessarily likely to be discharged. Therefore, we believe the approach we have taken to identify (possibly multiple) barriers is critical to highlight for potential users how their interventions might help a patient progress toward discharge.

#### Is the prescribed barrier the most likely to be resolved?

To conclude our discussion, we also checked whether the prescribed barriers were the ones with the highest resolution likelihood for that patient. Overall, 82.3% of actionable patients (above threshold 0.5) have a single prescribed barrier; among these patients, 89.4% are prescribed the barrier with the highest resolution likelihood (9.4% the second most likely, and 1.2% the third or lower barrier by likelihood). This suggests that it is necessary to consider both barrier resolution and its effect on discharge (as opposed to resolution alone) in any heuristic approach to solving BtD. Lastly, we note that if we instead consider all feasible patients (i.e., *J*_min_ = 0), then the percent with a single barrier is 37.6% (and among these, the prescribed barrier is the most likely 73.9% of the time).

### 3.4 Contribution of Personalization

To better understand the value of personalization, we evaluated its performance relative to a simpler approach. Namely, we replaced the personalized likelihoods with the baseline (aggregate) resolution likelihood for each barrier (based on the percent resolved by discharge in the training sample); as such, for a given barrier, we gave every observation with that barrier open the same estimated resolution likelihood. Using these, we reran the BtD algorithm; to allow a fair comparison, we selected that value of 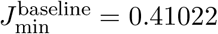 so that the number of actionable patients at this threshold was the same as the choice of 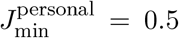 in the original, personalized approach, i.e., 1073 patient-day observations.

In Table 4, we compare the two models with respect to the retrospective performance metrics used earlier. In addition, we also considered how the choice of discharge time horizon impacts the criteria above. To do this, we created relaxed versions of the criteria, labeled *M* 2_next_ and *M* 3_next_, to be evaluated over a longer horizon, namely, whether a patient is discharged on the same day *or next day* following barrier resolution. Because some barriers (like facility acceptance) are more likely to have a day lag between resolution and when discharge is feasible (e.g., given need to arrange transport to the facility), *M* 3_next_ is likely more practically relevant.

**Table 4:** Comparison of the performance of BtD using personalized versus baseline likelihoods.

| Metric | Personal-<br>ized | Base-<br>line | $p$<br>value |
| --- | --- | --- | --- |
| ($M1$): All prescribed barriers resolved | 79.7% | 70.0% | < 0.001 |
| ($M2$): Discharged on the same day as prescribed barrier resolution,<br>among those with criterion $M1$ met | 54.3% | 55.7% | 0.581 |
| ($M3$): Both $M1$ and $M2$ met | 43.3% | 39.0% | 0.048 |
| ($M2_{\text{next}}$): Discharged on the same day or next day as prescribed barrier<br>resolution, among those with criterion $M1$ met | 82.4% | 82.3% | 1.0 |
| ($M3_{\text{next}}$): Both $M1$ and $M2_{\text{next}}$ met | 65.6% | 57.6% | < 0.001 |

We observe that personalized BtD significantly outperforms the baseline in terms of the metrics *M* 1 and *M* 3, meaning a higher percentage of patients have their prescribed barriers resolved, and a higher percentage have both their barriers resolved and are discharged shortly following that occurrence (statistical comparison are shown per a Fisher exact test). Importantly, we also see there is no appreciable difference in the likelihood of patients to be discharged within 24 hours *given that the prescribed barriers are resolved*, suggesting that the incremental improvement we see in the percent of patients meeting *M* 3 (the most important criterion in this setting) is attributable to the personalization of resolution likelihood (whereas *M* 2 reflects the discharge likelihood model’s performance, which one would expect to be similar).

The relative improvement in *M* 3 for personalization relative to baseline is 9.2% (95% CI [0.2%, 18.7%]). In comparison, the relative value of personalization is larger for *M* 3_next_ with a relative percent increase of 18.9% (95% CI [8.7%, 30.4%]). This is particularly important as the same-or-next-day approach is likely more practically reasonable. One of the reasons we believe that personalization is important in this context is the underlying patient heterogeneity noted earlier. A barrier that must be resolved in certain groups might not have a similar behavior in others, and therefore using additional covariate information is necessary to distinguish such variation in behavior observed in clinical practice.

We close our discussion of personalization by noting that while we have focused on the comparison of a personalized approach relative to a baseline which uses aggregate likelihoods, many approaches to barrier identification do not incorporate *any* resolution likelihood information whatsoever. For example, Safavi et al. (2019) display barriers which, if resolved, increase the discharge prediction any amount (i.e., if *J*_*d*_(**x**′) − *J*_*d*_(**x**^*c*^) *>* 0, where **x**′ reflects a barrier change from **x**^*c*^); but they do not incorporate the likelihood of resolution. Lastly, for a discussion of results across all percentages of actionable patients, see SM3.

### 3.5 Computational Efficiency

We conclude our assessment of the performance of the BtD algorithm by considering its computational efficiency. In the test set, the average time to solve the BtD algorithm per patient-day observation is 0.77 seconds, or an average total of roughly two minutes total time (139.6 seconds) per day (if the BtD algorithm is run sequentially; minimum by day of 30.8 seconds, maximum 193.3). This is well within the solution time required by the application. Importantly, the computational time required for BtD algorithm does not display any scaling behavior in the number of open barriers (Table SM3), in contrast to an enumerative approach.

#### Applicability to other model classes

We end our discussion of the computational results by noting that our approach can also be applied in settings where the underlying predictive model *J*_*d*_ is not a feed-forward neural network. In particular, our approach requires being able to (within a practically acceptable time limit) solve the problem of maximizing *J*_*d*_. Similar reformulation machinery as developed by Anderson et al. (2020) has also been used in the settings where *J*_*d*_ is from different model classes. Examples of this include the following: tree-based-models (Biggs et al., 2022); logistic regression (a special case of our approach); and black-box models (through model approximation). Note that more sophisticated model classes for *J*_*r*_ (than the regularized logistic regression used here) can be easily incorporated into our approach with no increase in complexity as these likelihoods are inputs to the BtD algorithm.

## 4 Model Robustness and Patient Prioritization

In this section, we turn our attention to two additional aspects related to our modeling approach: an analysis of model robustness (and a related extension); and an illustrated example use case for patient prioritization of post-hospital facility acceptance.

### 4.1 Model Robustness

To assess the robustness of the proposed model, we performed a variety of sensitivity analyses. Here we focus on *prediction uncertainty*, namely, how do prescriptions change with errors in the discharge model? We also formulate, solve, and empirically evaluate a robust optimization variant of the model to directly incorporate model uncertainty. (For a discussion of how the prediction horizon of 24-hours compares with a 48-hour prediction horizon, see SM5.)

To assess changes for a given patient prescribed a set *S* of barriers in a nominal model *M* with an alternative prescribed set *S*′ per model *M* ′, we use three different metrics:

1. *Differences in barriers:* we evaluate |*S* ∪ *S*′| − |*S* ∩ *S*′|, or the number of barriers different between *S* and *S*′ (we focus on the percentage of differences of 0 [i.e., equality], 1, or 2).
2. *Feasibility concordance*: if the models *M* and *M* ′ are in agreement about whether a patient is feasible, i.e., 𝟙(|*S*| *>* 0) = 𝟙 (|*S*′| *>* 0).
3. *Jaccard index*: we compute the Jaccard similarity index |*S* ∩ *S*′|*/*|*S* ∪ *S*′|.

#### 4.1.1 Changes in prescriptions given model estimation error

Our approach thus far has used the discharge prediction model (for *J*_*d*_) to create point estimates of discharge likelihood given that certain barriers are resolved. In reality, however, there is uncertainty in the estimated model, and it is possible that model uncertainty could translate into different prescriptions. Indeed, a disadvantage of most common neural networks is that, despite being efficient point predictors, they do not provide out-of-the-box output distributions that can be used to quantify prediction uncertainty. Gal and Ghahramani (2016) propose a tractable method called Monte Carlo Dropout (MCD) for quantifying uncertainty in predictions of neural networks. The method relies on the use of dropout regularization (Srivastava et al., 2014).

To study how the MIN set changes with variation in the discharge prediction model, we apply the MCD technique. To proceed, first we train a neural network with a 20% dropout rate (the choice of 20% dropout rate was based on a common choice in the literature, e.g., Gal and Ghahramani, 2016). We subsequently use this dropout-trained discharge prediction model (which we call the “nominal” model) to see how the prescriptions change when different versions of this dropout model are used. Specifically, for each observation in the test set, we perform the following 1,000 times: randomly silence 20% of the hidden nodes in the nominal model to create a new function 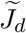 (in other words, we use a modified version of the nominal model to define 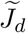); and solve the BtD problem on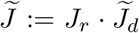 to identify a MIN set 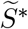 and the composite objective 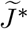.Then for each observation, we compute the Jaccard index relative to the nominal, non-silenced model (averaged across 1,000 runs); the percentage of runs in which 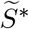 equals the nominal MIN set *S*^*^(or differs by at most 1 or 2 barriers); and the standard deviation of 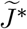 as a function of the initial discharge score.

The medians (across all observations) of these measures are shown in Figure 3 (distribution in Figure SM8; distribution of standard deviations of 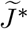 in Figure SM9). We observe that *for most patients* there is reasonably good agreement with the nominal model. For example, the median Jaccard index (across all patients feasible in the nominal model, i.e., prescribed at least one barrier) is 0.861 (IQR [0.801,0.960]), and it is consistent across the number of nominally prescribed barriers. However, for some patients, the prescriptions clearly differ across the 1,000 samples, and even differences of one barrier could be practically meaningful whenever it is the one of specific interest (especially for patients with fewer barriers in the MIN set per the nominal model). This suggests it might be necessary to address model uncertainty directly.

**Figure 3:**
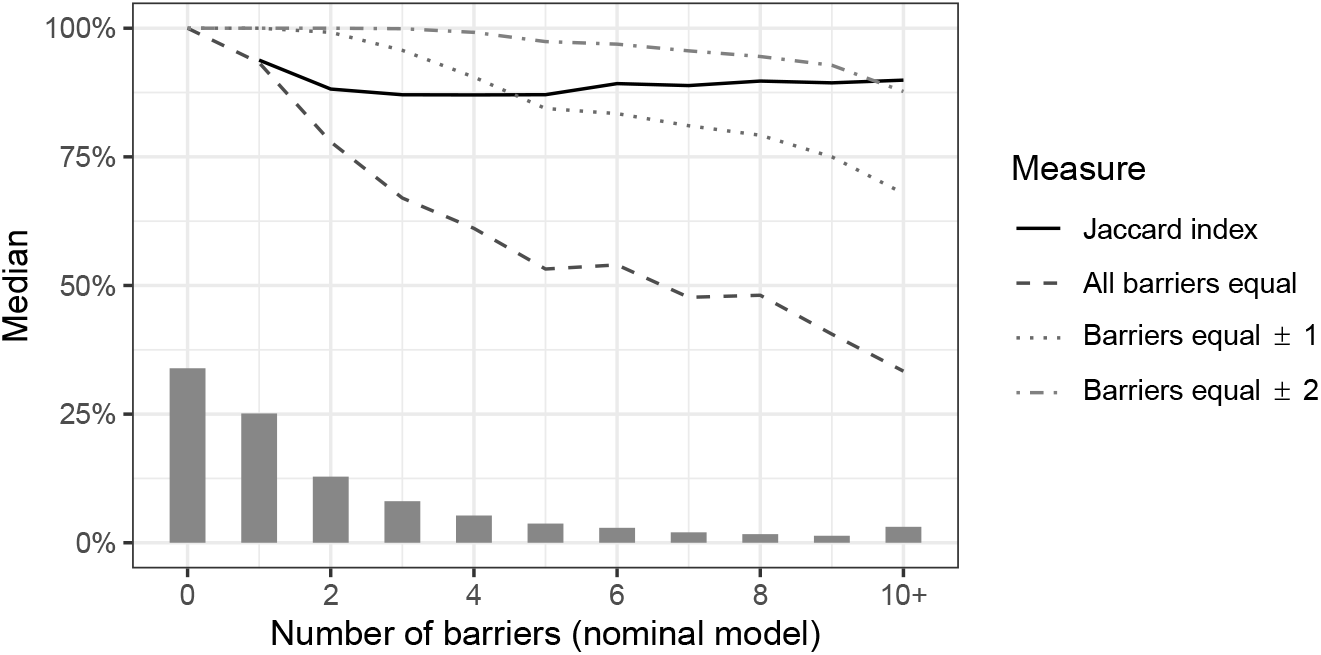
The median (across all test set observations) of Jaccard index and equality measures (computed across 1,000 simulated dropout runs). Observations are grouped by the number of barriers prescribed in the nominal (non-silenced) dropout model. The histogram shows the number of observations for each specified number of barriers. Observations with more than 10 prescribed barriers are grouped with 10.

#### 4.1.2 Model extension—Worst-case dropout prediction

We proceed by doing precisely that, namely, incorporating model uncertainty for the discharge prediction into the BtD optimization framework itself. To do so, we consider a robust optimization perspective, replacing *J*_*d*_ with a robust *worst-case* analogue *J*_*d,R*(*δ*)_ defined as the estimated discharge likelihood when a percent *δ* of hidden nodes are adversarially silenced (*δ* = 0% corresponds to the nominal model). To proceed, let us revisit the nominal value of *J*_*d*_, namely,

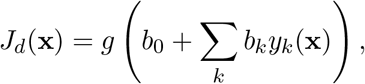

where *g* is the logistic function, 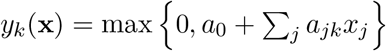,and **a** and **b** are coefficients from the dropout-trained model above. If a percentage *δ* ∈ [0, 1] of hidden nodes are silenced (say, corresponding to indices *D* ⊆ *{*1, 2, …, *h}*, with |*D*| = *δ · h*, where *h* is the number of nodes in the hidden layer), this objective with appropriate reweighting become

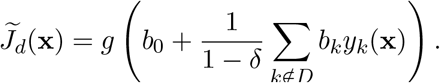

(If *D* is chosen randomly, this corresponds to the approach in the previous section.) Based on this setup, a worst-case (adversarial) silencing coincidences with finding the set of nodes that minimizes 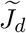.This can be written as

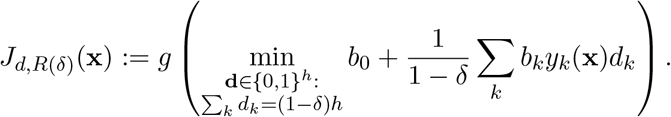

Using the standard duality trick, we can create a robust analogue of the BtD algorithm with *J*_*d*_ replaced by its *δ*-worst-case analogue *J*_*d,R*(*δ*)_ (see SM6). As with other typical robust optimization models, its structure is comparable to the nominal model.

To understand the performance of this alternative approach, we evaluate the same key metrics as in Table 3: above a given threshold *J*_min_, the percentage of patients meeting criteria *M* 1–*M* 3 and the corresponding values of 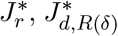,and 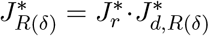 Instead of comparing as a function of *J*_min_, we use the percentage of actionable patients for a more direct comparison (as was done when evaluating personalization in Section 3.4). Given that a percentage actionable below 5% is the focus in Section 3, we restrict our attention to this range (see Figure 4; for the full range, see Figure SM5). The first observation is that the values of *M* 1–*M* 3, reflecting actual observed model performance, are generally consistent (though not identical) across different dropout percentages. However, the values of 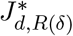 are lower with increasing *δ*, as one would expect—the model incorporates pessimism in the estimated discharge likelihood (in contrast, 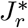 is generally invariant). Even for a small choice of *δ* = 3%, the values of 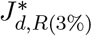 are nearly half of the corresponding values of 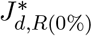 for a given percent of actionable patients.

**Figure 4:**
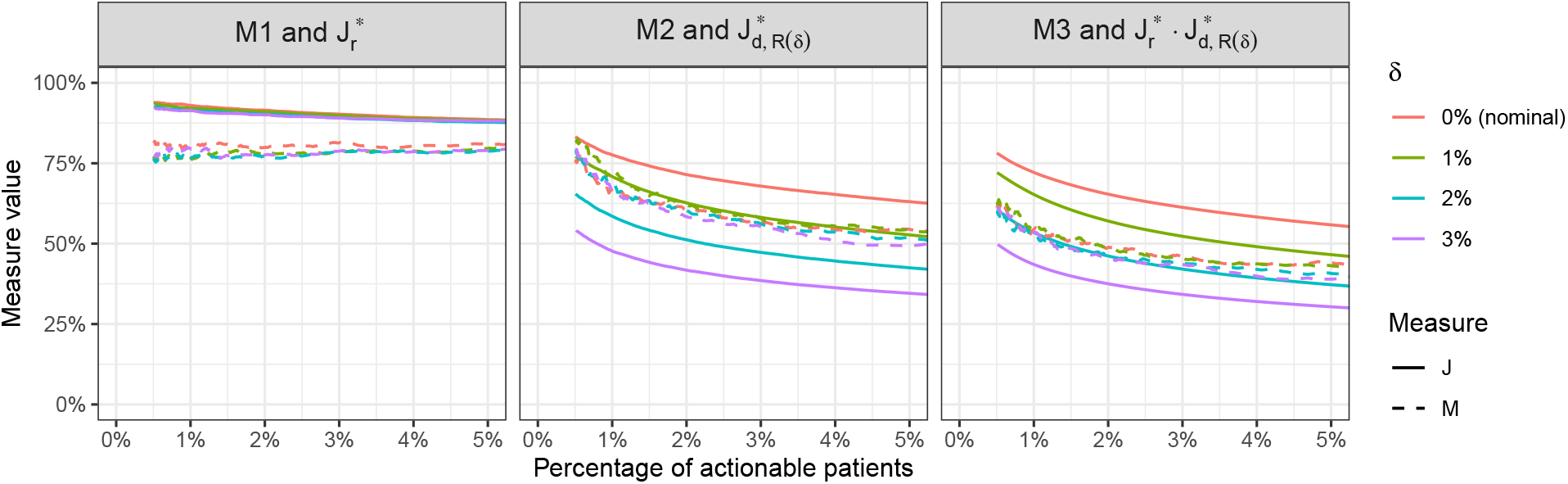
The values of M1–M3 (percentages) and 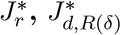,and 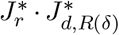 (averages) as a function of the percent of actionable patients for test set observations. *δ* denotes the adversarial dropout percentage.Actionable percentages below 0.5% are not shown given the small sample sizes.

In terms of calibration, given a desired choice for percent of actionable patients, *δ* = 2% gives excellent agreement between *M* 3 and 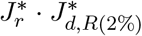,suggesting that the robust analogue of *J* ^*^ can be used as a means of achieving calibration in addition to addressing variance in the discharge prediction estimates. However, it is important to note that this calibration is not uniform across all possible choices of percent of actionable patients (Figure SM5). Nevertheless, given that the focus in our setting is on identifying a small number of actionable patients (on the order of 3-5%), *δ* = 2% appears to be a reasonable choice to achieve calibration if that is the desired objective.

Most importantly, we conclude our discussion of the worst-case dropout model by comparing how the actual *prescriptions* change as a function of *δ* as opposed to the objective values alone. Summary statistics about feasibility concordance and differences in the prescribed barrier set are shown in Figure SM6. The robust models tend to have more infeasible observations primarily due to observations which were previously feasible. Importantly, there are notable differences in the prescriptions, even for *δ* = 1%. For example, 13.7% of observations have a different prescription in 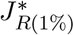 than in *J* ^*^ (due to both changes in feasibility and differences in which barriers are selected).

In other words, the robust model yields different MIN sets for some patients and does not only result in a modified objective function value.

We also evaluated how specific barrier prescriptions changed as a function of *δ* by focusing on the five barriers with the largest absolute change in number of times prescribed (across all observations) for the nominal problem compared with *δ* = 10% (Figure SM6). It is apparent that the robust model prescribes the “low urine output” barrier significantly fewer times (for *δ* = 2%, a 15.5% relative reduction, from 10.4% of observations to 8.8%). Interestingly, the robust model prescribes awaiting facility acceptance to fewer patients as well. We believe that the robust dropout model is itself of independent interest and should be the focus of future study. In particular, it would be critical to study prospectively (in an actual implementation) how reliably prescriptions can be used for the robust and nominal approaches (to discern practical value of this modification).

### 4.2 Patient prioritization—The case of post-discharge facility acceptance

One important practical application of the proposed prescriptive framework is to utilize it for informing barrier-specific interventions that require coordination of scarce resources at the hospital level. Here we focus on how the tool might be used specifically for the barrier of “awaiting post-hospital facility placement” in which the care team has identified that a patient requires a facility (such as skilled nursing facility, rehabilitation hospital, or long-term acute care) and needs to be accepted by one. Even though many hospitals are investing in partnerships with post-acute facilities (or creating their own facilities), they often lack insight into the operational workflow needed to identify which patients (across an entire hospital) should be prioritized for placement. At the same time, this process relies on manual escalations in cases where post-hospital facility bed need is one of the barriers significantly delaying a patient’s discharge. We focus on how a hospital could use our approach to identify patients to prioritize considering all patients awaiting a facility.

One way of using the BtD approach for this problem is as follows (for a given day):

1. First, we filter the patients for whom BtD prescribed the facility acceptance barrier. Let 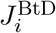 be the score obtained from the BtD algorithm for the *i*th patient.
2. Next, among these patients, we run a modified version of the BtD algorithm (“BtD-NoFacility”) which finds the *next best prescription* that does not contain the facility acceptance barrier. Let 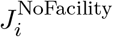 be the score obtained from the BtD-NoFacility algorithm for the *i*th patient.
3. Finally, we mark as “prioritized” the patients for whom the BtD score is above a threshold 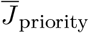,but the BtD-NoFacility score is below that threshold, i.e., 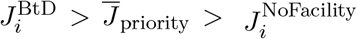. The threshold 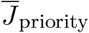 can be adjusted (calibrated) to increase or decrease the number of patients to be prioritized. (We tested a variety of other strategies and found similar results.)

We performed this experiment for the test sample of 153 days. There were 4523 patient-day observations where facility barrier was among the prescribed barriers (16.3% of all 27,601 test observations, or 30 patients a day). To best reflect actual use, we did this in a sequential manner (day by day), ensuring that each patient appears in the prioritized list at most once (namely, the first time a patient satisfies the prioritization rule described above). Similarly, we included a patient in the non-prioritized list only once (again, only the first time the patient does not satisfy the prioritization rule). A patient can appear in both lists. We set 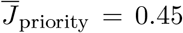 to yield an average of one patient per day to mark as prioritized.

Table 5 compares various metrics between prioritized and non-prioritized observations. We see that the percentage of patients for whom the facility barrier is resolved by the time of discharge is slightly higher in the prioritized group between prioritized and non-prioritized observations (88.3% versus 82.7%, though the difference is not statistically significant). Implicitly, prioritized patients have relatively high initial BtD score 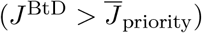,which could only be possible if their facility barrier had a high resolution likelihood.

**Table 5:** Using BtD to prioritize patients for facility barrier resolution (*J* _priority_ = 0.45). One outlier (present in both groups) with a long length of stay was excluded from calculating averages in (5) and (6).

| Metric | Prioritized | Non-Prioritized |
| --- | --- | --- |
| (1) Number of patients | 153 | 804 |
| (2) Number (%) of patients where facility barrier is resolved prior to discharge | 135 (88.3%) | 665 (82.7%) |
| (3) Number (%) of patients where the patient is discharged on the same or next day that the facility barrier is resolved, given that facility barrier is resolved | 121 (89.6%) | 505 (75.9%) |
| (4) % with facility barrier resolved and discharged on the same or next day | 79.1% | 62.8% |
| (5) Mean number of days between resolution and discharge | 0.44 | 1.36 |
| (6) Mean number of days between prescription and resolution | 2.60 | 5.95 |
| (7) Number (%) of patients with facility as the only prescribed barrier | 146 (95.4%) | 303 (37.7%) |

On the other hand, the percentage of patients who are discharged on the same day or next day after the facility barrier is resolved (given that it is indeed resolved) is significantly higher for prioritized patients compared to non-prioritized patients (89.6% versus 75.9%). Consequently, across all patients, the percent of patients who have their facility barrier resolved and are discharged within one calendar day is 79.1% of prioritized versus 62.8% of non-prioritized. This is the primary metric we use to evaluate the performance of the proposed prioritization policy (and is the facility-specific analogue of the metric *M* 3_next_ discussed in depth earlier).

We also observe that the proximity of facility barrier resolution to discharge, which is noticeably worse for non-prioritized patients than prioritized patients (1.36 days on average for resolution to discharge versus 0.44 days), lends support to the notion that the facility barrier was not the singular barrier preventing discharge for non-prioritized patients (otherwise one would expect discharge to follow shortly after resolution). This partially addresses a consistent concern raised in discussions with clinical stakeholders, namely, that prioritization of barriers (for the stated purpose of timely discharge) which does not actually result in a patient being discharge-ready can damage trust in any prioritization process. Importantly, prioritized patients very frequently have facility acceptance as their only prescribed barrier (95.4% versus 37.7% for non-prioritized), suggesting that the prioritization criteria is identifying patients with fewer barriers even though this was not explicitly included in the logic.

We close our discussion by noting the potential benefit of this prioritization scheme. The average time from being marked as prioritized to barrier resolution (among those with the barrier resolved) is 2.60 days, equating to a total of 354 bed-days in the test period (or an average daily census of 2.3 patients). This represents the potential benefit of such an intervention if this time interval could be shortened. While the effort required to shorten this interval is highly nontrivial, we believe that it is helpful to contextualize the possible bed-days saved if such an effort was pursued (in this case, from prioritizing an average of one patient per day).

## 5 Conclusion

As hospitals face increased congestion, timely discharges have become a pillar of efficient patient flow. However, central capacity planners have limited visibility into how to make targeted, small-scale interventions for patients who are most likely to get discharged within the next day. In this study, we combine machine learning and discrete optimization models to identify a small subset of barriers. To the best of our knowledge, this is the first time that this modeling approach is applied in healthcare. Our experiments on real data corroborate (i) the utility of the proposed objective function, (ii) the efficacy of the BtD algorithm, (iii) the value of personalization, and finally (iv) how the model can be adapted including a robust analogue which can better calibrate probability estimates and an example use case.

Future work includes adding capacity constraints to our models. For example, some barriers in our model correspond to those with a “global” capacity constraint such as the number of available post-discharge facility beds or slots in an MRI machine in a given day. Adding such between-patient constraints raises a variety of important methodological questions such as scalability of various decomposition approaches. In addition, while our approach focused on using barrier resolution likelihood as the balancing measure for discharge likelihood, one could easily replace this with a more direct measure of “cost” required to resolve a barrier (such as relative time or effort required to resolve a barrier). We suspect that even in such cases, incorporating information about resolution likelihood (perhaps as a function of effort expended) will be crucial, and additional empirical work is needed to appropriately parametrize such general models.

## Data Availability

No data is publicly available.

## Supplemental Material for

This Supplemental Material contains the following additional tables, figures, and analysis for completeness:

1. features included in the data (Table SM1);
2. the full list of 149 barriers;
3. information on performance of different classifier algorithms for discharge prediction (Table SM2);
4. discussion of model calibration and corresponding calibration curves for the discharge prediction score (Figure SM1);
5. the barrier resolution model training process;
6. pairwise comparison of test set AUC, percentage of observations with a given barrier, and percentage resolved for that barrier;
7. details of the formulation of the problem of maximizing *J*_*r*_ subject to lower bounds on *J*_*d*_ as a MIP by using the approach of Anderson et al. (2020);
8. the performance of *J* as a function of the number of selected barriers;
9. additional data on the relative value of personalization in the BtD algorithm (Figure SM4);
10. solve times for the BtD algorithm as a function of the number of open barriers;
11. the percentage of time each barrier is prescribed (given that it is open) in the 24-hour and 48-hour models;
12. the distribution of metrics (Jaccard index, set agreement percentages, and standard deviation of 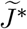)in the 1,000 simulated dropout models; and
13. the worst-case dropout estimation formulation and results.

## SM1. Additional predictive modeling details

### 1.1 List of barriers

The list of 149 barriers (by thematic grouping) is as follows:

1. Bedside measurements: Cognition; High Pain; Level of consciousness; Orientation level; Speech
2. Bloodwork: Cryoprecipitate; Plasma; Platelets; RBC
3. Awaiting cardiac procedures: Cath; EP; Echocardiogram
4. Case management: Bed Hold; Discharge Services; High Risk
5. Consult services: Addiction Services; Allergy; Anesthesiology; Cardiology; Dentist; Dermatology; Endocrinology; Gastroenterology; General Surgery; Gerontology; Gynecology; Hematology; Infectious Diseases; Internal Medicine; Interventional Radiology; Nephrology; Neurology; Neurosurgery; Nutrition Services; Oncology; Ophthalmology; Oral Maxillofacial Surgery; Orthopedic Surgery; Otolaryngology; Pain Management; Palliative Care; Physical Medicine Rehab; Plastic Surgery; Podiatry; Psychiatry; Pulmonology; Pyschology; Rheumatology; Smoking Cessation Program; Social Work; Surgery: Burn; Surgery: Neuro Spine; Surgery: Ortho Hand; Surgery: Ortho Spine; Surgery: Trauma and Acute Care; Thoracic Surgery; Urology; Vascular Surgery
6. Intake/output: Irregular diet; No oral intake; Emesis; Not passing flatus; Low Urine output
7. Post-acute placement: await facility acceptance; await home services acceptance
8. Imaging: CT; MRI; Other; US; XR
9. Labs: CRE - high; CRE - increased; GLU - high; GLU - low; HGB - decreased; HGB - low; INR - high; K - high; K - low; Lactate - high; NA - high; NA - low; Troponin - increased; WBC - high
10. Lines, drains, and airways: BiliaryDrain; ChestTube; EFMS; EpiduralCatheter; GiTube; NegPressureWoundTherapy; OpenSurgicalWound; PICC; PenroseDrain; SuctionDrain; SuprapubicCatheter; UrinaryCatheter
11. Medications: Antiemetic; Antipsychotic; Colonoscopy Prep; Diuretic; Heparin; IV Antibiotics; IV Fluid; IV Narcotic; Nebulized Meds; Nitroglycerin; PO Narcotic; Phenobarbital; Steroids; TPN; Tamsulosin; Tube feeds; Warfarin
12. Microbiology pending: Anaerobic; Blood; CSF; Fluid (not CSF); Fluid Culture Smear (not CSF); Fungal; Mycobacteria; Stool; Tissue; Urine; Wound
13. Occupational therapy: Final dispo recommendation
14. Physical therapy: Activity; Final dispo recommendation; Sit to Stand Level of Assistance; Soft restraint L wrist; Soft restraint R wrist; Stand to Sit Device Used; Stand to Sit Level of Assistance; Supine to Sit Level of Assistance; Surface to Surface Device Used; Ambulation Assistance
15. Speech language pathology: Clinical Swallow Pharyngeal Phase; Clinical Swallow Response; Diet recommendation; Final dispo recommendation; Medication recommendation; Swallowing recommendation
16. Vital signs: BP; Cardiac rhythm; O2 Device; Pulse; Respiration; SpO2; Stool occurrence; Temperature

**Table SM1:**
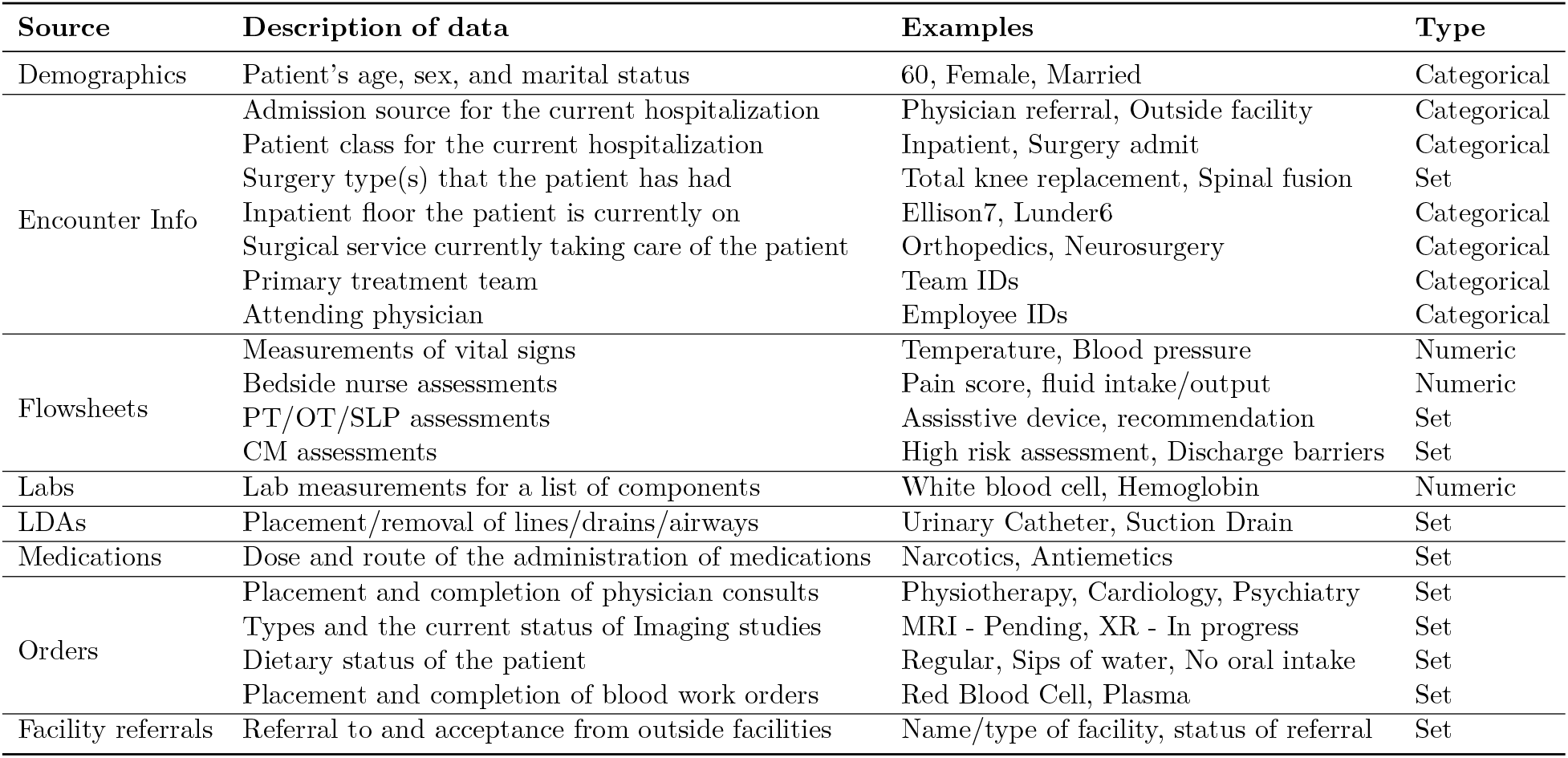
Overview of the data sources used. “Categorical” indicates that only one of the values in a given list can be used for each patient, while several values can be used if the type is “Set.”

**Table SM2:** Performances of different classifier algorithms on the test sample

| Prediction horizon | Classifier | AUC | Recall | Precision |
| --- | --- | --- | --- | --- |
| 24 hours | Classification Tree | 0.811 | 0.545 | 0.521 |
|  | Logistic Regression | 0.862 | 0.620 | 0.581 |
|  | Neural Network | 0.877 | 0.619 | 0.611 |
| 48 hours | Neural network | 0.871 | 0.740 | 0.727 |

### 1.2 Out-of-sample prediction performance

For precision and recall calculations in Table SM2, we used the *unbiased* threshold score above which a patient is predicted to be discharged and below which is predicted not to be discharged. We defined the unbiased threshold as that which would equally balance model recall and precision in the training set. Moreover, when this threshold is used to convert prediction scores to binary classifications, the number of predicted discharges in a given subset is expected to be equal to the number of actual discharges in that subset, hence the name “unbiased threshold.” For the 48-hour model (SM4 below), we only evaluated the neural network model.

### 1.3 Model calibration

For a predictive model *h* : *F* → [0, 1], where *F* is the feature space and the observed outcome data is a binary *y* ∈ *{*0, 1*}*, model calibration requires assessing how well *h*(**x**) estimates ℙ(*y* = 1|**x**), i.e., the probability that *y* = 1 given feature vector **x** ∈ *F*. There are numerous approaches to assessing model calibration. For our purposes, we follow one of the common approaches based on the *Brier score* for *h*. For a set of data 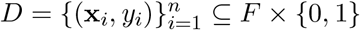,it is defined as

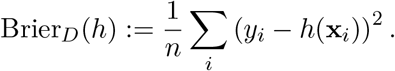

Lower Brier scores typically correspond to better calibration for a given prediction task.

In order to assess whether a given model *h* is properly calibrated on a data set *D*, we apply the Spiegelhalter *Z*-test (Huang et al., 2020). In particular, a model is defined as calibrated per a Spiegelhalter test (at the 0.05 level) if Spieg_*D*_(*h*) ≤ 1.96, where

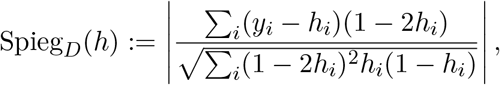

and *h*_*i*_ := *h*(**x**_*i*_). We chose to apply this definition because it is easy to use in settings where many models are being tested and where it is not feasible to visually inspect usual calibration plots; indeed, this is important in the subsequent section on barrier resolution models. (Note that Spiegelhalter tests are sometimes interpreted via *p* values instead. In particular, a *p* value can be found by computing *p* = 2 *·* Φ (−Spieg_*D*_(*h*)), where Φ is the cumulative density function of a standard normal; therefore, calibration corresponds precisely with failing to the reject the null at a significance level of 0.05.)

Calibration curves for the 24-hour discharge prediction model are shown in Figure SM1. The prediction model consistently overestimates discharge likelihood across all values of the scores. Therefore, we applied Platt scaling (estimated based on the training sample) to the final discharge predictions to obtain well-calibrated discharge probability estimates (*p*-value for Spiegelhalter test of 0.082, 0.004, and 0.228 on the training, validation, and test sets, respectively). The original Brier scores were 0.139, 0.143, and 0.145; and the final Brier scores were 0.109, 0.111, and 0.113.

**Figure SM1:**
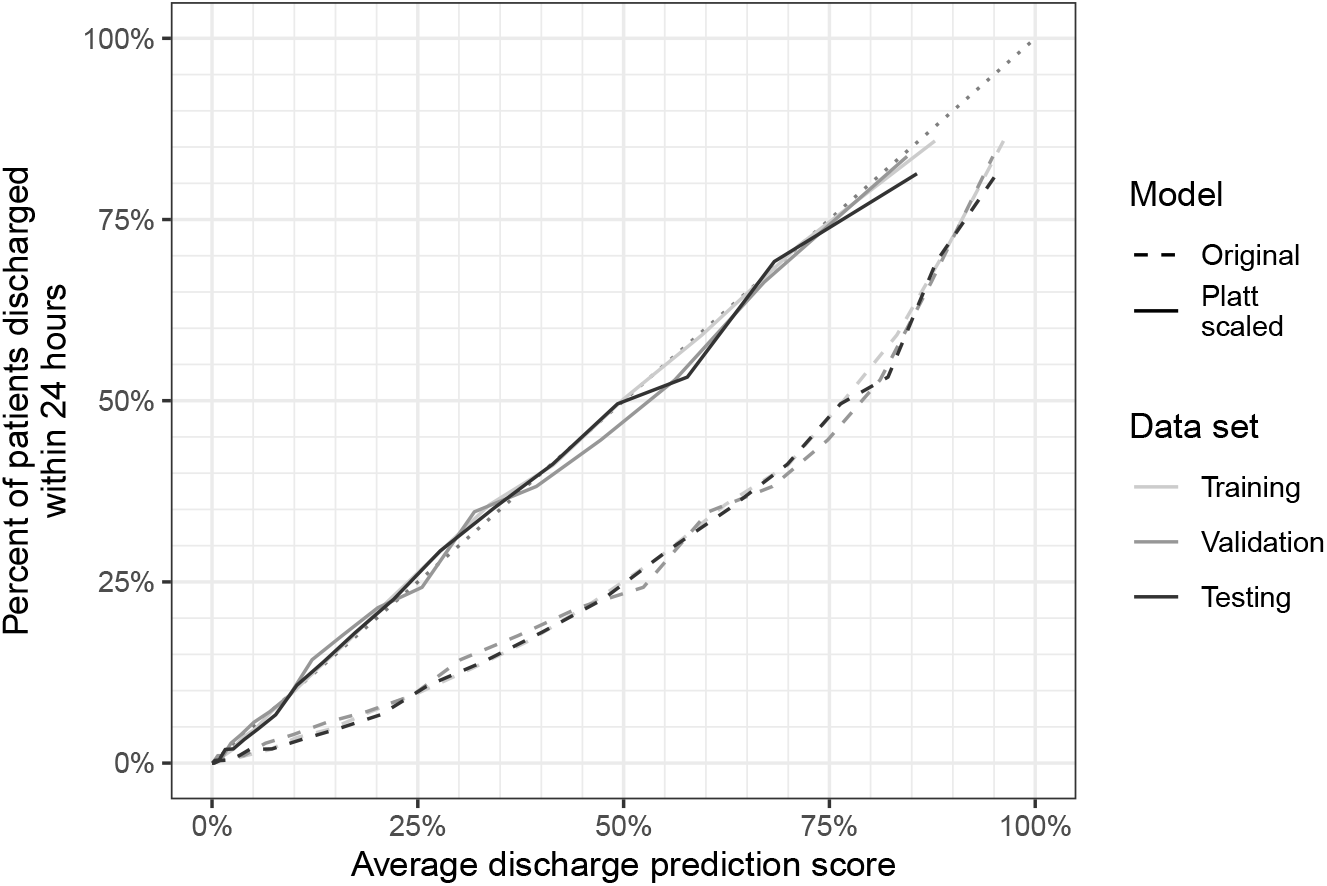
Calibration curves for the discharge prediction scores on the three data sets. Predictions grouped into 20 equally sized groups for each of the data sets. Two models are shown: the original, and the version after Platt scaling based on the training set. A dotted line on the diagonal (perfect calibration) is included for reference.

### 1.4 Resolution likelihood model training

To train barrier resolution models, for each of the 149 barriers we performed the following:

1. On the subset of the training data with that barrier open, we train a sequence of *f*_1_-regularized logistic regression models. In particular, we use `glmnet` (Friedman et al., 2010) to compute the regularization path for regularization parameters *λ*_1_ *> λ*_2_ *> · · · > λ*_min_ *>* 0 (we use the default automated grid choice and remove any *f*_2_ regularization). Using the parameter *λ*_1se_ corresponding to the parameter for the model within one standard error of that with the lowest cross-validation error (as performed only based on the training set), we exclude any models with *λ < λ*_1se_. (The *λ*_1se_ choice is based on standard practice recommendations.) This results in a set of regularized logistic regression models 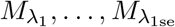.We assume that we have *K* such models (i.e., |*{λ*_1_, …, *λ*_1se_*}*| = *K*).
2. We also train *K* ordinary logistic regression models 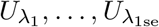 defined as follows: for each model *M*_*λ*_ on the regularization path, we identify the set of nonzero variables for that model and train an ordinary (unregularized) logistic regression model (again with outcome being whether the barrier is resolved) but only on the subset of features for which the corresponding coefficient in *M*_*λ*_ is nonzero. (The motivation for training such unregularized models as well in this setting is that regularization is known to potentially adversely affect model calibration.)
3. For the set of 2*K* logistic regression models, we exclude any models with validation AUC below 0.5; we also exclude any models which fail to calibrate per a Spiegelhalter test (either on the training data or on the validation data). Among the remaining models, we pick the one that maximizes AUC_val_ − Brier_val_ − max{AUC_train_ − AUC_val_, 0}, where AUC_*D*_ denotes the AUC on data set *D*. The motivation for this choice is that we simultaneously want to maximize validation AUC while minimizing Brier score and controlling the imbalance between the training and validation AUCs.

Based on this process, there were 116 barriers with validation-calibrated models. There were 25 barriers for which there are no calibrated logistic regression models with validation AUC above 0.5; for such barriers, we use the baseline resolution percentage (in the training and validation sets combined) for all observations. There were also 8 barriers which were never present in the training set, never present in the validation, or never resolved in both; resolution likelihoods were set uniformly to 0 for these barriers.

### 1.5 Resolution likelihood model performance

In Figure SM2, we show the performance of the resolution likelihood models on the test set for the subset of 116 barriers for which personalized likelihoods are used (the remaining 33 use the baseline resolution percentage from the combined training and validation sets per the validation procedure described in the main text). The performance measure shown is AUC, and we consider the relationship between AUC and the underlying percentage of observations with each barrier as well as the percent of observations with the barrier resolved (among those with that barrier). Each point corresponds to a barrier. Note that five barriers have test AUC strictly below 0.5 (although all training and validation AUCs were strictly larger than 0.5 by design; these three barriers correspond to those triggered in fewer than 2% of observations). In general, we see that there is a significant positive correlation between AUC and the percent of observations, as noted in the main text.

**Figure SM2:**
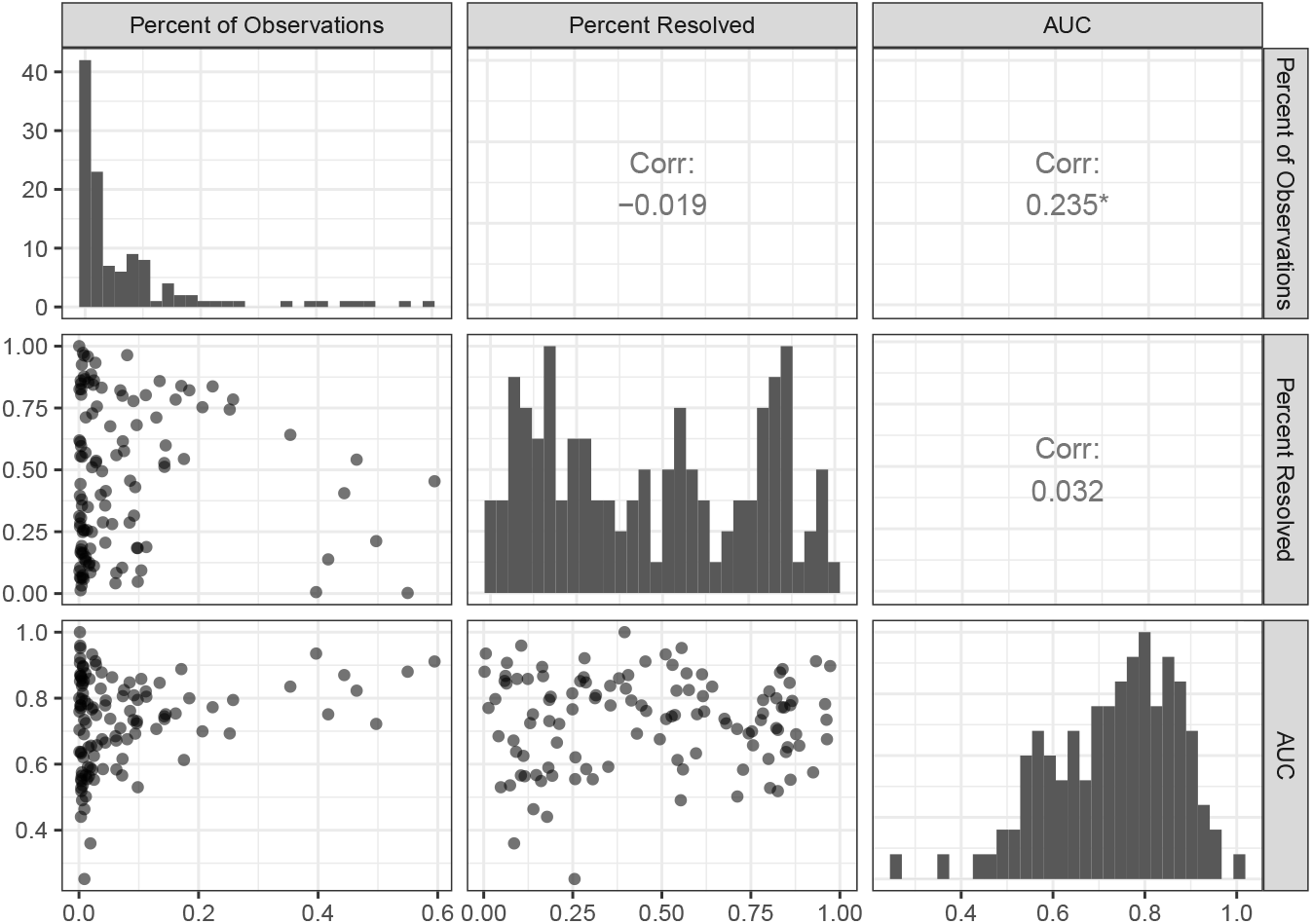
Resolution likelihood models—Pairwise comparison of test set AUC, percent of observations with a given barrier, and percent resolved for that barrier. The diagonal shows histograms for each of those three measures; the upper diagonal displays correlation coefficients; and the lower diagonal shows scatter plots.

### 1.6 Performance of *J* as function of number of selected barriers

In Section 4, we assessed the performance of *J* for a randomly selected subset of up to five barriers (choosing from one through five with equal probability) to reflect a variety of scenarios including a single barrier being selected up to five barriers. (If there were fewer than five barriers, we sampled at most the number of open barriers.) To better understand how the performance of *J* varies as a function of the number of barriers, we also consider when *k* open barriers are selected, for *k* ∈ *{*1, 2, …, 10*}*. In particular, for each *k*, we consider the subset of test set observations with at least *k* open barriers; for each observation, we randomly select a subset of *k* of those barriers, and then compute the corresponding value of *J*_*r*_, *J*_*d*_, and *J*, along with the label as defined in Section 3.

For each choice of *k*, we compare several metrics:

- Brier score (relative to the labeled outcome);
- AUC for *J* (relative to the labeled outcome);
- AUC for *J*_*r*_ (relative to all-barriers-resolved outcome); and
- AUC for *J*_*d*_ (relative to the discharged-same-day outcome, only on the subset of patients with all selected barriers resolved).

The corresponding results are shown in Figure SM3. Note that the Brier score for the labeled outcome (whether patient has all selected barriers resolved *and* is discharged within one day of all such barriers being resolved) decreases as the number of barriers increases. At the same time, the overall AUC for *J* generally increases, going from 0.799 for *k* = 1 to 0.946 for *k* = 10. Interestingly, the AUC for *J*_*d*_ is generally decreasing, while the AUC for *J*_*r*_ is increasing. One possible explanation for the decrease in AUC for *J*_*d*_ is that with a larger number of selected barriers, more time is likely required to pass before barrier resolution (and hence the discharge model, which does not account for changes in non-barrier features, degrades in performance). Note that the increasing width of the *J*_*d*_ confidence intervals is because this is calculated on an increasingly small subset of observations.

**Figure SM3:**
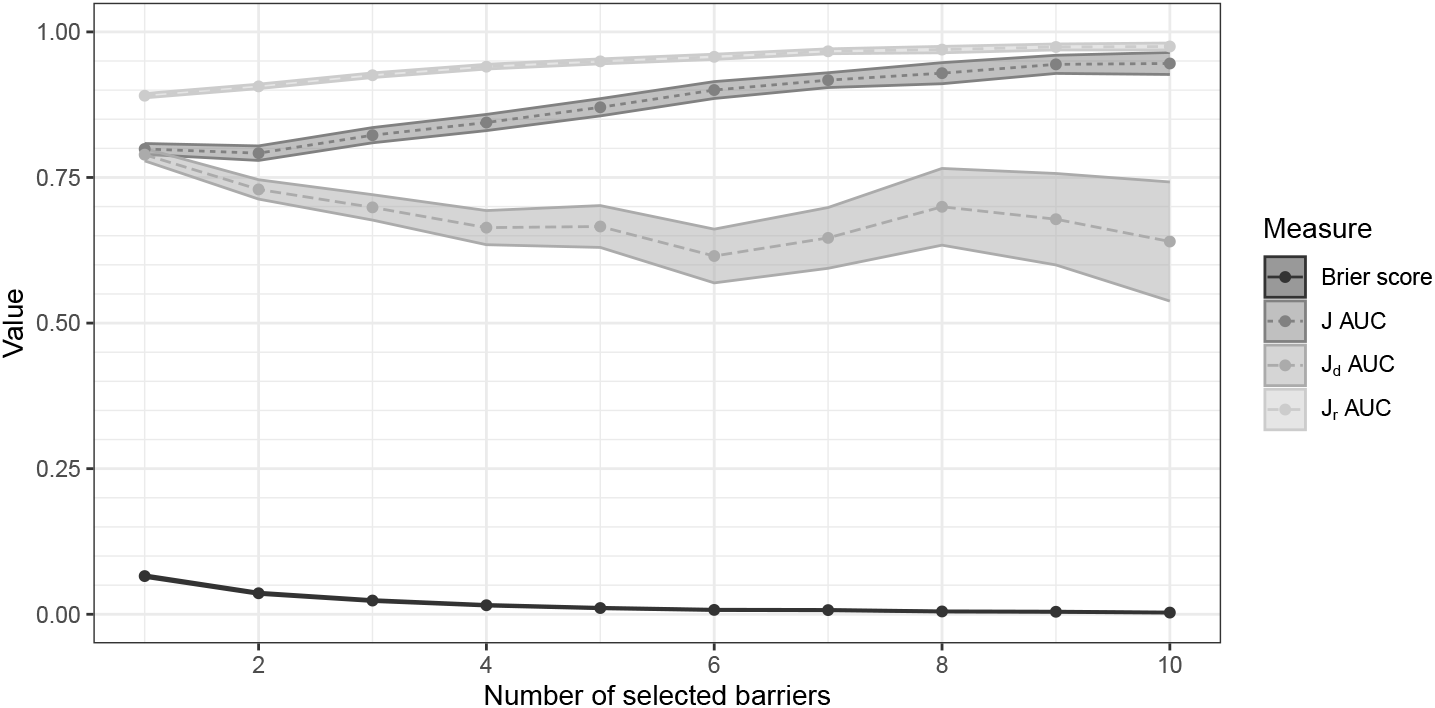
Brier score and AUC metrics for composite objective as a function of the number of barriers selected (95% CIs are shown for each metric). The Brier score shown is for *J* (not *J*_*d*_ nor *J*_*r*_).

## SM2. MIP formulation approach

Here we show the details of formulating the problem of maximizing *J*_*r*_ subject to a lower bound on *J*_*d*_ can be formulated as a linear mixed integer program (MIP), thereby making use of the formulation approach taken in Anderson et al. (2020). For expository purposes, we begin by examining the structure of a single-hidden-layer neural network to illustrate how the prediction score is calculated (the discharge prediction model as used in the main text has this structure).

Figure SM4 shows an example of a (feed-forward) neural network with 5 input nodes, 3 hidden nodes and a single output node. In this mock example, **x** = (*x*_1_, *x*_2_, *x*_3_, *x*_4_, *x*_5_) is the input feature vector. The values *a*_*j*1_, *a*_*j*2_, *a*_*j*3_ are the coefficients of the *j*^*th*^ feature in the first, second, and third hidden nodes, respectively. Moreover, *a*_01_, *a*_02_, *a*_03_ are the biases added to the the first, second, and third hidden nodes, respectively. The outputs of hidden nodes (*y*_1_, *y*_2_, *y*_3_) are calculated as follows: 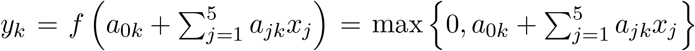.Similarly, *b*_1_, *b*_2_, *b*_3_ are the coefficients of the first, second, and third hidden nodes in the output node, and *b*_0_ is the bias added to the output node. The output prediction score *z* is calculated as follows: 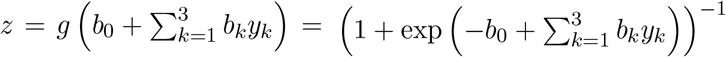.

**Figure SM4:**
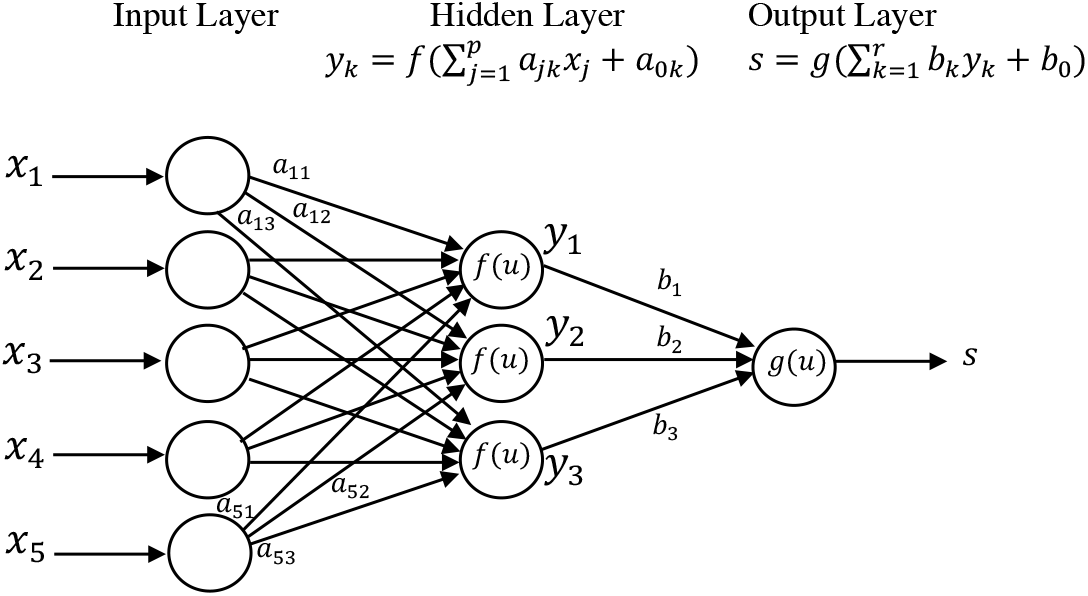
Network structure and parameters for an example feedforward neural network classifier

To formalize the structure illustrated in Figure SM4 and adapt it to our problem setting, we partition the feature space into 4 components as described in Section 2 in the main text 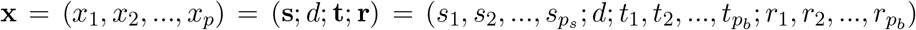. In other words, we define 4 sets of indices *S* = *{*1, 2, …, *p*_*s*_*}, 𝒟* = *{p*_*s*_ + 1*}, 𝒯* = *{p*_*s*_ + 2, …, *p*_*s*_ + 1 + *p*_*b*_*}*, and *ℛ* = *{p*_*s*_ + 1 + *p*_*b*_ + 1, …, *p*_*s*_ + 1 + 2*p*_*b*_*}* that represent the set of indices of the static, days since admission, trigger and resolution features, respectively, in our feature space. In Section 3.1 in the main text, the trained neural network model had *p* = *p*_*s*_ + 1 + 2*p*_*b*_ input nodes, *h* hidden nodes (on a single layer), and a single output node to predict the discharge likelihood within 24 hours for a given feature vector 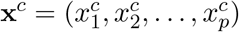 of size *p*. The following are parameters of the prediction model:

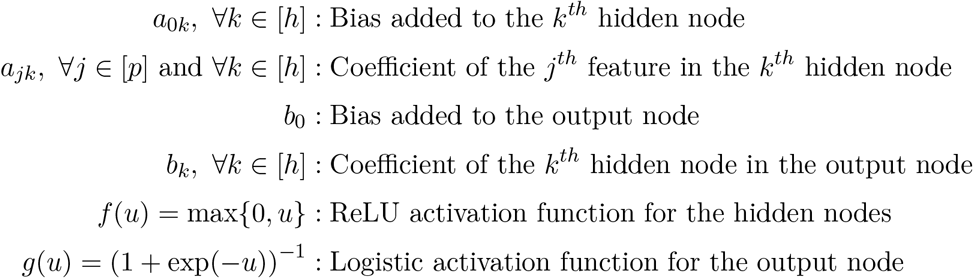

Next, we introduce three sets of decision variables to express the problem as a MIP:

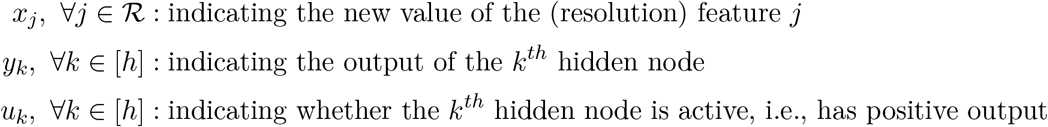

For a given feature vector and parameters, the output *y*_*k*_ of the *k*^*th*^ hidden layer can be written as follows:

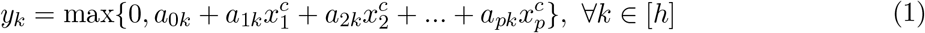

which can be expressed linearly with the following sets of constraints and auxiliary binary variables *u*_*k*_.

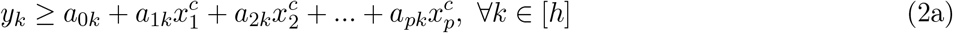

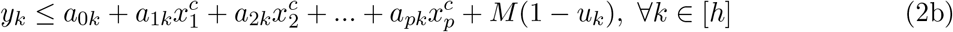

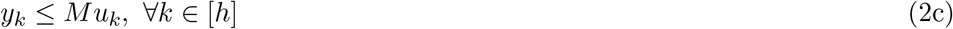

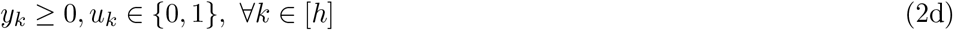

(Here, *M* is an appropriately calibrated constant.) Similarly, the prediction score *z*, which is the output of the output layer, can be written as

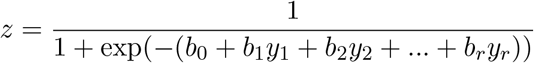

Note that if we would like this modified score to be greater than a pre-determined threshold *z*_0_, then this can be written as a linear constraint:

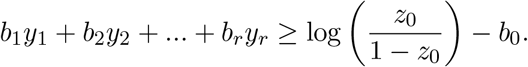

Finally, we incorporate the restrictions on the allowed changes to the feature vector. Specifically,

- *x*_*j*_ ∈ *{*0, 1*}*, ∀*j* ∈ *ℛ*: we are only allowed to change the barrier resolution features from 0 to 1
- 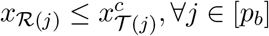: only a currently open barrier can be resolved,
- 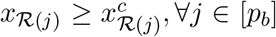: a barrier that is already resolved cannot be reverted back to being unresolved

where *ℛ* (*j*) and *𝒯* (*j*) indicate the *j*^*th*^ element of the index set *ℛ* and *𝒯*, respectively.

Note that the current features **x**^*c*^ are fixed parameters in our subproblem. Similarly, the personalized resolution log-likelihoods log(*L*_*j*_) are parameters pre-calculated from the barrier resolution prediction models for each barrier. Finally, the coefficients of the neural network model **a** = (*a*_*jk*_), **b** = (*b*_*k*_) are also the fixed parameters in the subproblem. Given a threshold discharge prediction score *z*_0_, the following is the MIP formulation for the aforementioned subproblem, hereinafter referred to as SP(*z*_0_; **x**^*c*^):

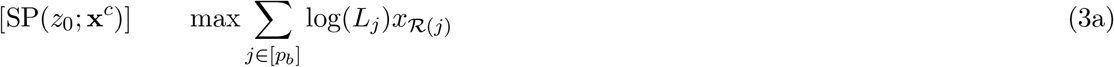

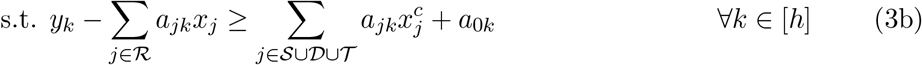

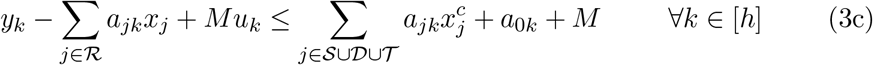

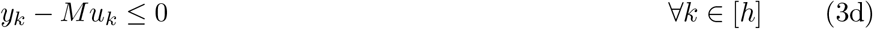

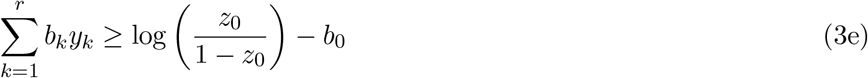

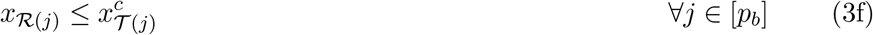

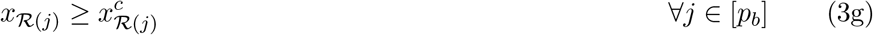

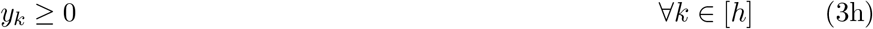

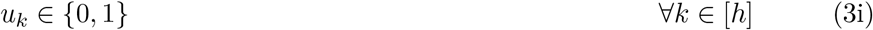

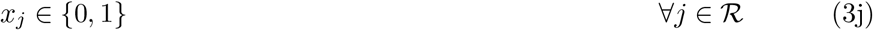

In the above formulation, constraints (3b)-(3d) linearize the ReLU activation function, i.e.,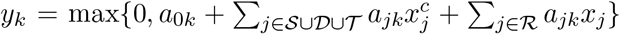. Constraint (3e) imposes the restriction that the modified score has to be above the pre-determined threshold *z*_0_. Constraints (3f) impose the condition that a barrier cannot be resolved if it is not triggered in the first place and (3g) impose that a barrier cannot be reverted back to being “unresolved” if it is currently resolved. Finally (3h)-(3j) define the types and bounds of the decision variables.

Note that the formulation contains 2*r* + *p*_*b*_ variables and 4*r* + 2*p*_*b*_ + 1 constraints. While the formulation explicitly includes variables *x*_*j*_ even for barriers *j* that are not triggered, a simple preprocessing step can eliminate these (as they will equal zero in any feasible solution). Because of the Platt scaling used, we replace *z*_0_ by the inverse of the appropriate Platt scaling function applied to *z*_0_. We set *c* = 0.01 for all analyses, equating to solving for *z*_0_ ∈ *{*0, 0.01, 0.02, …, 0.99*}*.

## SM3. Other actionability thresholds

We also evaluated the criteria *M* 3 and *M* 3_next_ across all possible percentages of actionable patients (the preceding results equate to 3.9% actionable). There are several notable observations (see Figure SM4):

1. The baseline model exhibits undesirable behavior for small percentages of actionability whereby the performance is *worse* when considering small subsets of patients. As having a small actionable percentage is precisely the regime of practical interest, this reinforces the importance of using the personalized approach.
2. For larger percentages of actionable patients, the personalized approach is indistinguishable from the baseline in terms of the criterion *M* 3. However, there are actionable percentages where the baseline model is not feasible, in contrast to the personalized model, suggesting that the baseline approach is overly restrictive when using aggregated likelihood values.
3. For the same-or-next-day criterion *M* 3_next_, the personalized approach is uniformly superior to the baseline. Given the points raised about this metric above, the personalized approach is clearly preferable.

**Figure SM4:**
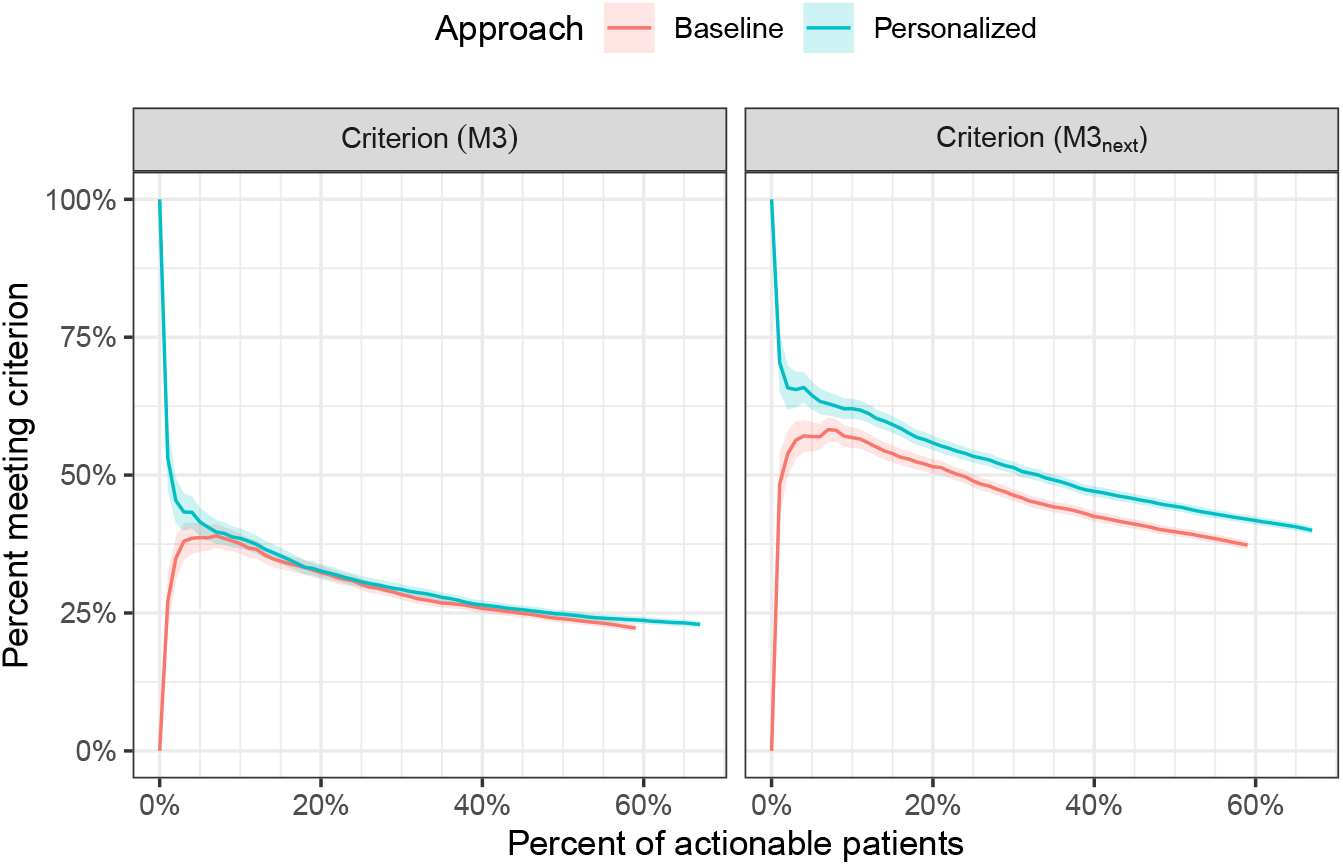
Comparison of the percent of patients meeting criteria *M* 3 and *M* 3_next_ as a function of the percent of patients who are actionable from the personalized versus aggregated approaches (generalization of Table 4).

## SM4. Computations

### 4.1 Solve time for BtD algorithm

The average computation time for the BtD algorithm as applied to the test set is shown in Table SM3. We see that the average time is essentially independent of the number of open barriers (overall averaging less than 1 second), in contrast to an enumerative approach for which the computation time scales exponentially.

**Table S3:** Computational performance of BtD algorithm as a function of the number of open barriers. Computation time is an average (in seconds). A row for the total is shown at the bottom.

| # Open barriers | # Observations | Average time (s) |
| --- | --- | --- |
| $\leq 5$ | 5004 | 0.85 |
| 6 | 2414 | 0.88 |
| 7 | 2660 | 0.88 |
| 8 | 2669 | 0.86 |
| 9 | 2432 | 0.83 |
| 10 | 2069 | 0.80 |
| 11 | 1867 | 0.79 |
| 12 | 1540 | 0.77 |
| 13 | 1223 | 0.74 |
| 14 | 1034 | 0.67 |
| 15 | 854 | 0.66 |
| 16 | 686 | 0.59 |
| 17 | 567 | 0.56 |
| 18 | 488 | 0.48 |
| 19 | 395 | 0.50 |
| $\geq 20$ | 1699 | 0.38 |
| (Any) | 27601 | 0.77 |

## SM5. Alternative 48-hour model

To better understand the implications of the modeling choice of predicting discharge within 24 hours, we also created a prediction model for discharge within 48 hours (using the same setup as the 24-hour model, again with appropriate post-training calibration; see Table SM2). Given this new model, we solved the BtD algorithm using the 48-hour analogue of *J*_*d*_. To understand how prescriptions changed, we evaluated how concordant the two models are in terms of feasibility (as defined in Section 2.4), how the numbers of prescribed barriers compare, and the relationship between the prescriptions in the two models.

### Feasibility concordance

In terms of feasibility, the two models are in agreement 89.3% of the time (57.7% of the time the patient is feasible in both models, and 31.5% in neither); 9.7% feasible in the 24-hour model but not the 48-hour model; and 1.0% feasible in the 48-hour model but not the 24-hour model. In other words, typically barriers are *more likely* to be prescribed when assessing discharge likelihood over a *shorter* time horizon (although this is not uniformly true). Intuitively, this occurs because discharge scores are higher overall for a longer prediction horizon; therefore, higher baseline prediction scores mean that the ability to increase a score (through prescribing barriers) is smaller, and any such score increases must be balanced with the barrier resolution likelihood. Put a different way, prediction over a longer time horizon reduces the discriminative power of the BtD approach. To best appreciate this, it is easier to think about an extreme modification of the model, where we predict discharge within 100 days. In this case, all baseline discharge prediction scores would already be close to 1 and the model would rarely prescribe any barriers (unless the barrier had nearly 100% resolution likelihood).

### Number of barriers prescribed

The number of barriers prescribed in the 24-hour model tends to be the same (59.9%) or larger (33.9%), while the 48-hour model prescribes more barriers in 6.2% of instances. The average number prescribed in the 24-hour model is 2.0 barriers (standard deviation 2.6), while for the 48-hour model the average is 1.6 (2.3). Among the subset of feasible patients from each approach, the average is 3.0 barriers (2.6) in the 24-hour model versus 2.8 (2.5) in the 48-hour model. Across all patients (feasible and infeasible), the difference in number prescribed (24-hour minus 48-hour) averages 0.4 (SD 0.9; median of 0, interquartile range [0, 1]).

### Relationship between prescriptions

Given the above observations, we evaluated whether the 48-hour prescribed barriers are a subset of the 24-hour barriers. Indeed, this is the case for 87.7% of observations: 41.2% of the time, the 48-hour model is infeasible (i.e., no barriers exist which can increase *J*_*r*_*·J*_*d*_ from its initial value, hence none is prescribed) and therefore the prescribed set is tautologically a subset; and 46.4% there are at least some prescribed barriers in the 48-hour model and they are a subset of the 24-hour barriers. We also compared the prescription percentage (relative to number of times as an open barrier) between the two models; see Figure SM7. On an individual barrier level, the concordance between the prescriptions in the two models ranges from 92.6% to 100.0% (median 100.0%, IQR [99.8%, 100.0%]).

## SM6. Worst-case dropout prediction

We now turn our attention to a formulation of the BtD problem using the worst-case dropout prediction (Section 4.1.1). As defined in Section 4.1.2,

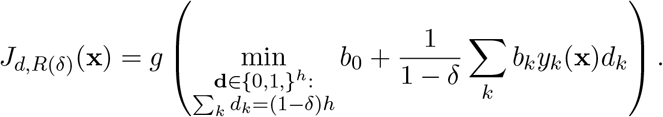

Using the standard duality trick, the BtD model with *J*_*d*_ replaced by its *δ*-worst-case analogue *J*_*d,R*(*δ*)_ yields the following variant of the subproblem SP(*z*_0_; **x**^*c*^), with additions or modifications to the nominal problem shown with a “*\**” to the left of the constraint:

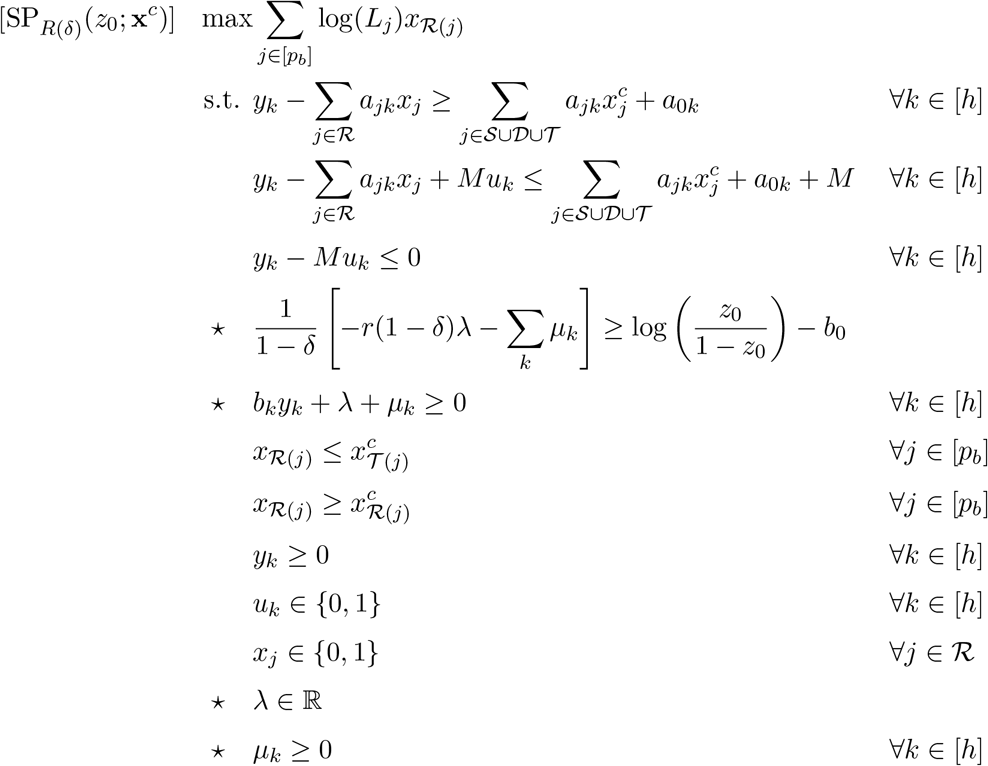

In other words, it has *h* + 1 additional variables (*λ* and ***µ***), 2*h* additional constraints, and one constraint modified. This is again a linear MIP which can be solved in the same way as the nominal problem. In the model used in the main text, *h* = 100, so *δ* = 0.05 would correspond to five silenced nodes, for example.

### Performance of 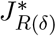 **as a function of** *δ*

In addition to the figure shown in the main text, we also compared the performance of the robust approach across different percentages of actionable patients. The corresponding results are shown in Figure SM5. We see that the agreement between *M* 3—the percent of patients with all prescribed barriers resolved and discharged on the same day as resolution—and 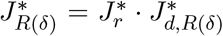 is different across different actionable percentages, suggesting that if calibration is desired for a specific percentage, the appropriate choice of *δ* might differ (for example, the nominal model is already reasonably effective at 40% actionable, and *δ* = 2% is effective for actionable thresholds below 5%). We also compared feasibility concordance, agreement in barrier set prescriptions, and changes in specific barrier prescriptions in the robust dropout model relative to the nominal model across thresholds *δ* ∈ *{*0%, 1%, …, 10%*}*—see Figure SM6 (note that while most values plateau by *δ* = 10%, they do continue changing with larger choices of *δ*, which is not shown here for ease of interpretation). For changes in specific barriers, we restricted our attention to the barriers with the five largest absolute changes in the number of times prescribed (and show percentage relative to the number of times prescribed in the nominal).

**Figure SM5:**
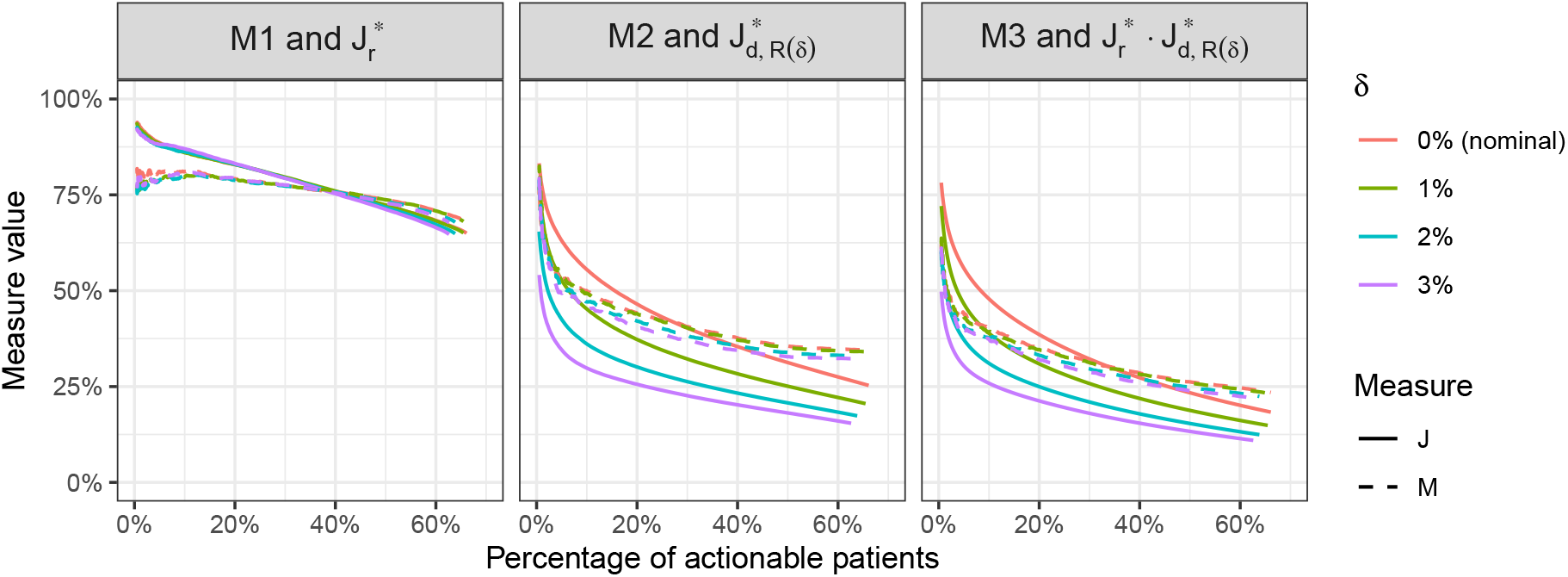
The values of *M* 1–*M* 3 (percentages) and 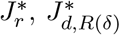,and 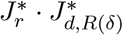 (averages) as a function of the percent of actionable patients for test set observations. *δ* denote the adversarial dropout percentage. Actionable percentages below 0.5% are not shown given the small sample sizes.

**Figure SM6:**
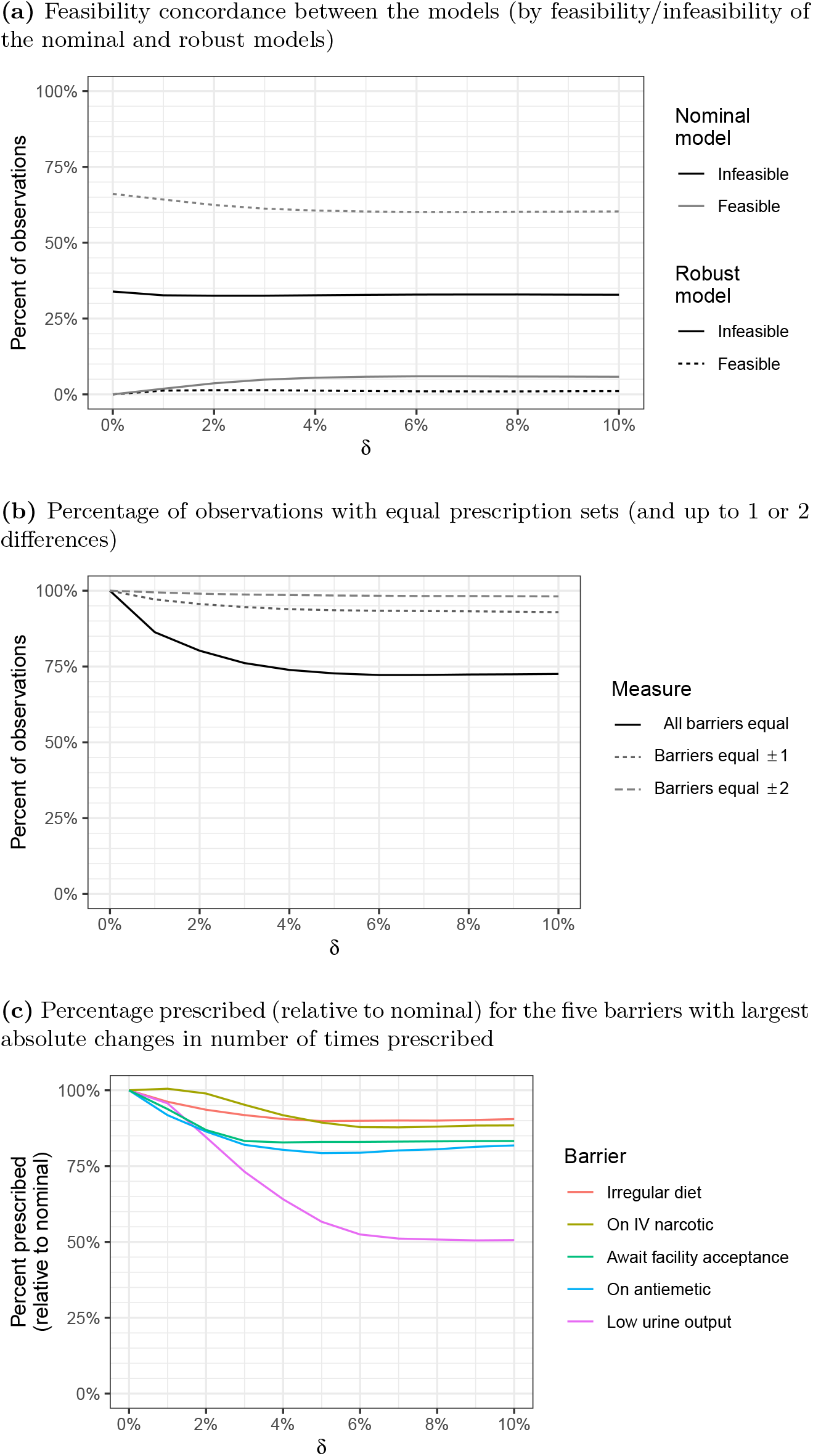
Feasibility concordance, agreement in barrier set prescriptions, and changes in specific barrier prescriptions in the robust dropout model relative to the nominal model across thresholds *δ*∈ *{*0%, 1%, …, 10%*}*. Note that *δ* = 0% is the nominal model. For changes in specific barriers, we restrict our attention to the barrier with the five largest absolute changes in the number of times prescribed (and present relative to number prescribed in the nominal).

**Figure SM7:**
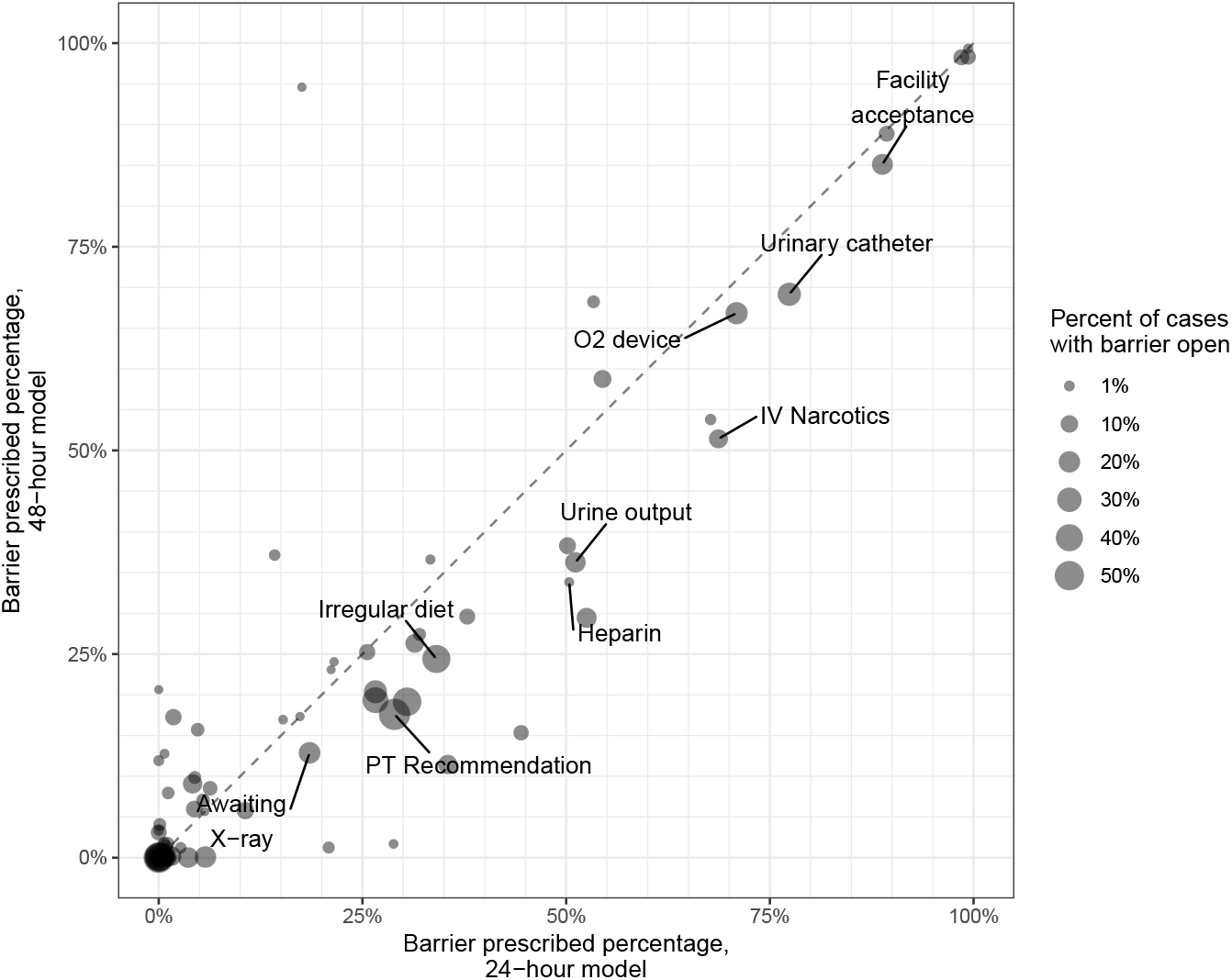
Comparison of the average barrier prescription percentage (among those with the barrier open) in the test set for the 24-hour and 48-hour models. Each point is a barrier and its size corresponds to the percent of patients with that barrier triggered. The same barriers as in the main text are labeled for reference (except “PO Narcotics”, removed for legibility). Abbreviations: PO=“medication taken by mouth”; IV=“intravenous”; PT=“physical therapy”; O2=“oxygen.”

**Figure SM8:**
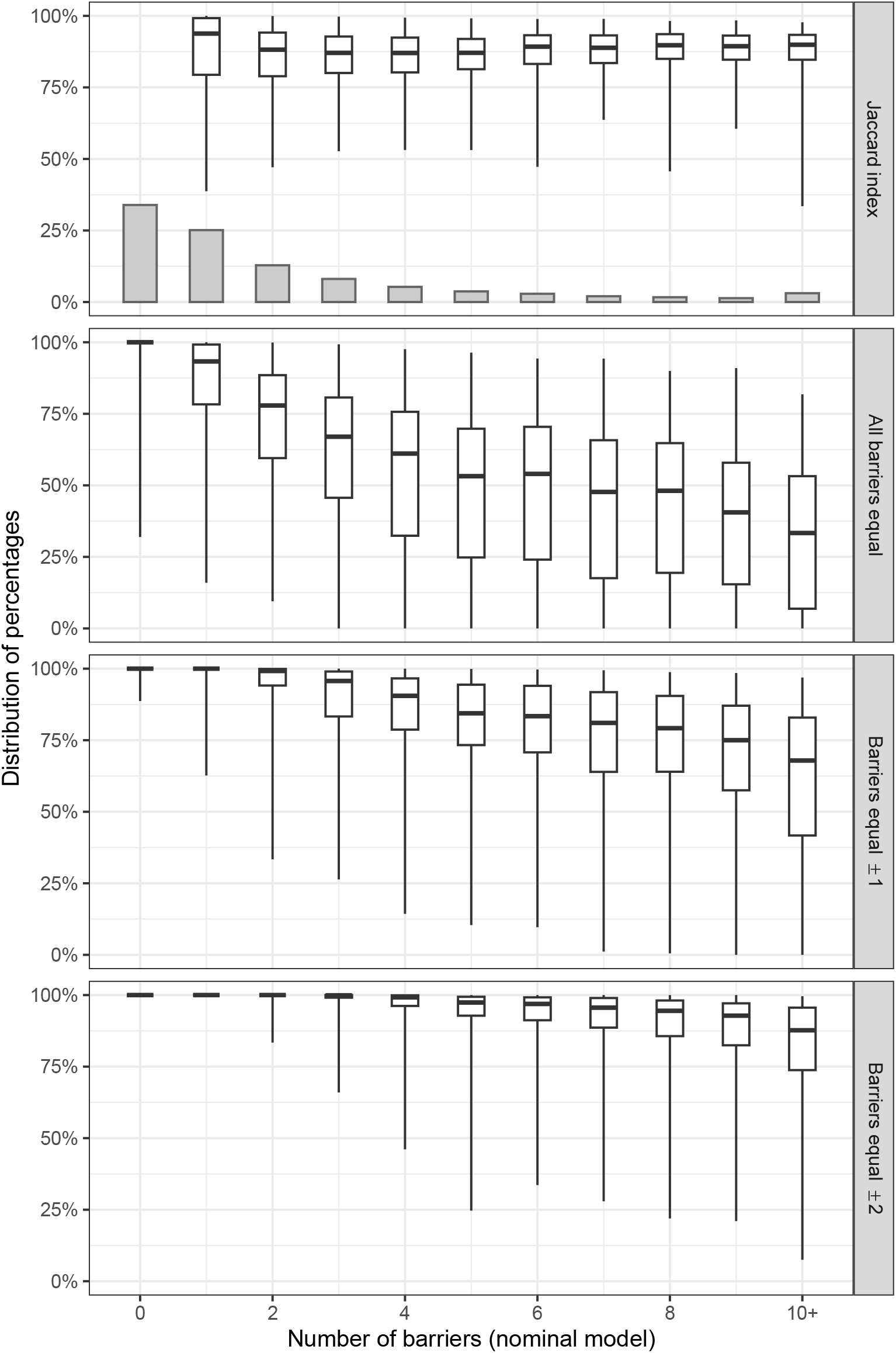
The distribution of measures (computed across the 1,000 simulated dropout runs) for all test set observations (medians reflect those reported in Figure 3). Observations are grouped by the number of barriers in the nominal (non-silenced) dropout model. The top plot shows a histogram reflecting the number of observations for each specified number of barriers. Observations with more than 10 prescribed barriers are grouped with 10. Whiskers extend to the 1st and 99th percentiles.

**Figure SM9:**
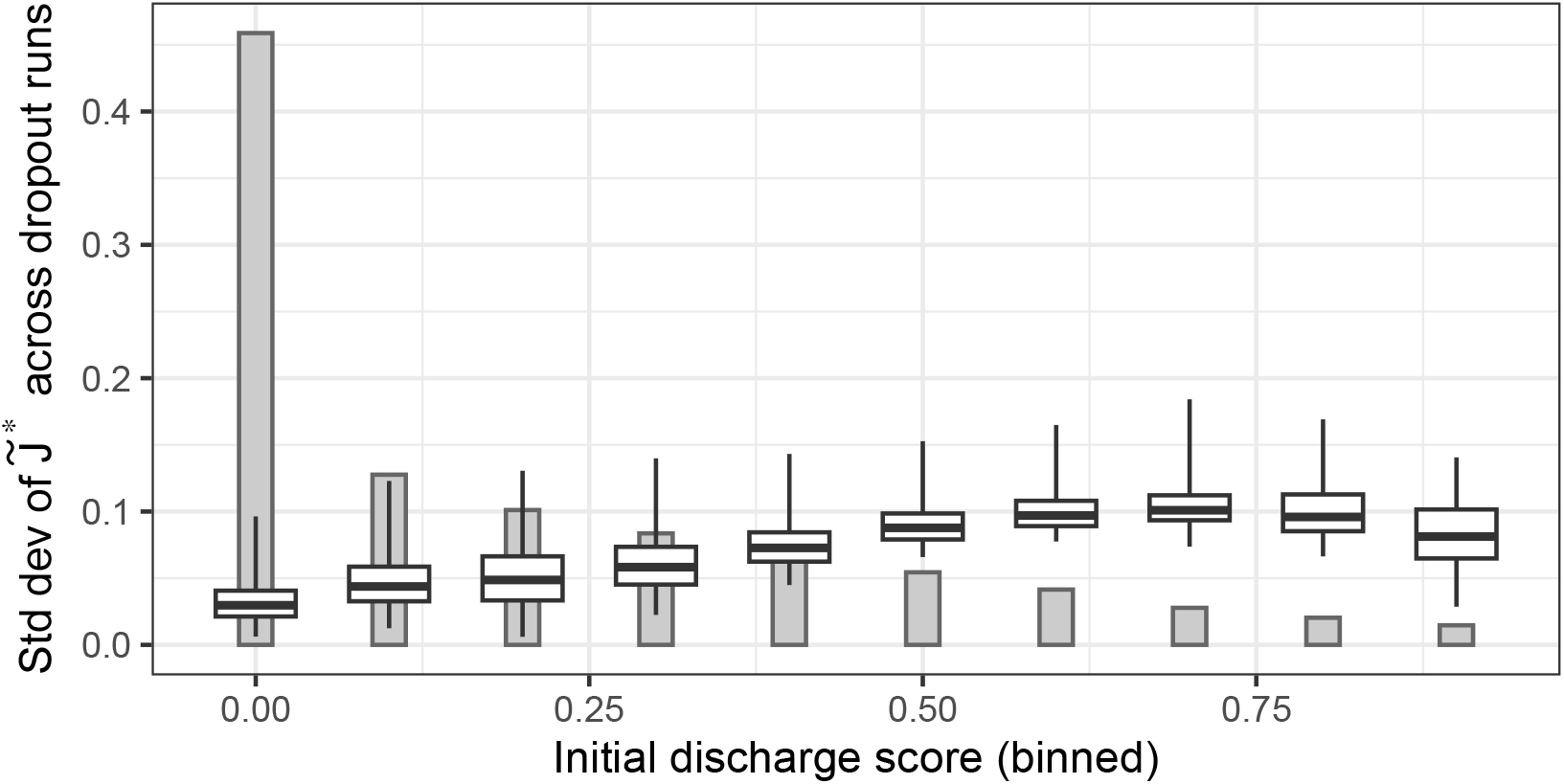
The distribution of the standard deviation of 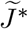 (as measured across the 1,000 simulated dropout runs). Observations are grouped into 10 evenly-spaced groups by initial discharge score in the dropout model ([0, 0.1), [0.1, 0.2), and so on). The histogram reflects the number of observations for each specified number of barriers. Whiskers extend to the 1st and 99th percentiles.

